# Prevalence of IgG and IgM to SARS-CoV-2 and other human coronaviruses in The Democratic Republic of Congo, Sierra Leone and Uganda: A Longitudinal Study

**DOI:** 10.1101/2023.03.08.23286979

**Authors:** Bolarinde J. Lawal, Katherine E Gallagher, Jonathan Kitonsa, Daniel Tindanbil, Kambale Kasonia, Abdoulie Drammeh, Brett Lowe, Daniel Mukadi-Bamuleka, Catriona Patterson, Brian Greenwood, Mohamed Samai, Bailah Leigh, Kevin K. A. Tetteh, Eugene Ruzagira, Deborah Watson-Jones, Hugo Kavunga-Membo

## Abstract

**Objectives:** We assessed the prevalence of immunoglobulin G (IgG) and immunoglobulin M (IgM) against four endemic human coronaviruses (HCoVs) and two SARS-CoV-2 antigens, among vaccinated and unvaccinated staff at health care centres in Uganda, Sierra Leone, and the Democratic Republic of Congo (DRC).

**Methods:** Government health facility staff who had patient contact in Goma (DRC), Kambia District (Sierra Leone), and Masaka District (Uganda) were enrolled. Questionnaires and blood samples were collected at three timepoints over four months. Blood samples were analysed with the Luminex MAGPIX®.

**Results:** Among unvaccinated participants, the prevalence of IgG/IgM antibodies against SARS-CoV-2 RBD or N-protein at enrolment was 70% in Goma (138/196), 89% in Kambia (112/126) and 89% in Masaka (190/213). IgG responses against endemic HCoVs at baseline were not associated with SAR-CoV-2 sero-acquisition during follow-up. Among vaccinated participants, those who had evidence of SARS-CoV-2 IgG/IgM at baseline tended to have higher IgG responses to vaccination compared to those SARS-CoV-2 seronegative at baseline, controlling for the time of sample collection since vaccination.

**Conclusions:** The high levels of natural immunity and hybrid immunity should be incorporated into both vaccination policy and prediction models of the impact of subsequent waves of infection in these settings.

## Introduction

Natural immunity and ‘hybrid immunity’, generated after a mixture of both natural infection and vaccination, could illicit more effective and longer-lasting protection against subsequent symptomatic infection with SARS-CoV-2, than vaccination alone [1, 2]. Vaccine coverage with at least one dose of a prophylactic COVID vaccine was 4.6% in the Democratic Republic of the Congo (DRC), 34.1% in Sierra Leone, and 40.5% in Uganda in August 2022[3]. The outcome of future waves of SARS-CoV-2 infection will be determined by both natural and hybrid immunity and whether other common circulating pathogens like endemic human coronaviruses (HCoVs) produce a non-specific protective response.

Prior to SARS-CoV-2, the most common human coronaviruses (HCoVs) were the alpha-HCoVs (OC43, HKU1), and the beta-HCoVs (NL63 and 229E), which cause mild to moderate respiratory tract diseases. Globally, most people acquire one or more of these viruses in their lifetime, and develop symptoms such as cough, sore throat, runny nose, fever, headache, and general malaise [4-6]. In pre-pandemic samples, OC43 and HKU1 anti-spike (anti-S) antibodies have displayed cross-binding to the SARS-CoV-2 Spike protein (S-Protein) with some neutralising activity[7, 8], although it is still unclear whether prior exposure to HCoVs provides any protection against SARS-CoV-2 infection [9, 10]. Additionally, both vaccination and natural infection with SARS-CoV-2 have been shown to generate IgG which binds S-proteins from other beta-HCoVs (HKU1 and OC43; but not alpha-HCoVs NL63 or 229E) [7, 9, 11, 12]. SARS-CoV-2 infection and/or vaccination may stimulate memory responses and generate production of IgG to HCoVs in adults with prior exposure to HCoVs or there may simply be cross-binding to common epitopes. Results differ on whether these anamnestic responses are protective against infection with SARS-CoV-2. Pre-existing IgG antibodies to the nucleocapsid protein (N-protein) of HCoV 229E were weakly correlated with remaining uninfected with SARs-CoV-2 infection in a cohort of health care workers (HCWs) but were not correlated with protection from infection in a large retrospective analysis of stored samples and electronic health records[13]. However, both studies found that pre-existing IgG antibodies to HCoVs correlated with less severe symptoms in those who did become SARS-CoV-2 positive[13, 14]. It may be important to account for levels of prior exposure to other HCoVs when estimating natural immunity to SARS-CoV-2 and to control for the presence of cross-reactive antibodies to HCoVs when using seroprevalence to SARS-CoV-2 as a marker of exposure to the virus[7, 8].

We aimed to assess the prevalence of IgG and IgM to two SARs-CoV-2 antigens, the nucleocapsid protein (NP), a marker of natural exposure, and the receptor-binding domain (RBD) of the spike protein which could be a marker of natural exposure or vaccine-induced immunity, among vaccinated and unvaccinated staff at health care centres in Uganda, Sierra Leone, and the DRC. We assessed IgG and IgM responses to four endemic HCoVs and explored whether these correlated with a reduced risk of acquisition of SARS-CoV-2 during the study or affected the immune responses to subsequent vaccination. We hypothesised that there may be different rates of sero-acquisition across the different settings that may be correlated with differences in HCoV seroprevalence or other risk factors.

## Methods

### Study Design

This was a longitudinal observational study of SARS-CoV-2 serology among staff working within primary healthcare facilities. Blood samples were collected over four months at three timepoints between February-June 2021 in DRC, March-July 2021 in Sierra Leone and July-November 2021 in Uganda.

### Study Setting and Population

The study took place in the city of Goma, DRC, and both urban and rural locations in Kambia District, Sierra Leone, and Masaka District in Uganda. A list of all government health facilities in each area was compiled. In Masaka District, all 25 government health centres were selected. In Goma, all 21 urban and accessible health centres were selected. In Kambia District, a random number generator was used to select 29 health facilities, proportional to the total number of health posts and health centres in the district.

Sierra Leone reported its first COVID-19 case on 31 March 2020 [15], the DRC on 10 March 2020 [16], and Uganda on 21 March 2020 [17]. COVID-19 vaccination was launched in Sierra Leone on 15 March 2021 [18], in DRC on 19 April 2021 [19], and in Uganda on 10 March 2021 [20, 21].

### Sample Collection & processing

We administered a short questionnaire to consenting staff at the selected health facilities to collect demographic data, information on comorbidities, vaccination status, reported symptoms of COVID-19 and contact with confirmed COVID-19 cases in their household, community or at work. At each site, a trained phlebotomist in appropriate Personal Protective Equipment (PPE), compliant with local guidelines, collected a 5ml venous blood sample into a Serum Sample Separator Tube. In each country, blood samples were allowed to clot upright and transported from the field to the local laboratories at 2-15°C. In the laboratory, serum was separated into aliquots and was frozen at −20°C. Each participant was followed up at two further visits at two-month intervals. At each visit, updated information on his/her COVID-19 vaccination status and contact data were recorded and a blood sample was collected. One aliquot of serum from each visit from DRC and Uganda was shipped at - 80°C to the research laboratory in Kambia town, Sierra Leone.

All samples from all participants were run on the Luminex MAGPIX® platform (Luminex, Texas) to test for IgG and IgM antibodies to SARS-CoV-2 NP and RBD, OC43 NP, 229E NP, HKU1 NP, NL63 S1 and MERS NP by adapting a previously established assay [22] (Patterson et al., 2022 [Manuscript in preparation]). The IgG assay achieved sensitivity and specificity of 98.8% and 97.9% for RBD, respectively, and 95.3% and 99.0% for NP, respectively. The IgM assay achieved sensitivities and specificities of 95.6% and 100% for RBD, respectively, and 65.8% and 100% for NP, respectively. (Patterson et al., 2022 [Manuscript in preparation]). The results of the multiplex assay were reported in units of Mean Fluorescent Intensity (MFI) and were defined as seropositive if they were 3 standard deviations above the mean MFI of 40 European pre-pandemic negative controls provided by Public Health England (Public Health England, 2016). Positive SARS-COV-2 control pools were from NIBSC and were used as plate controls (20/B770; 20/130).

### Statistical Analysis

Seroprevalence to SARS-CoV-2 was defined if a sample was seropositive for IgG or IgM to either the SARS-CoV-2 RBD or the NP and was tabulated stratified by vaccination status. Seroprevalence to other endemic HCoVs was not possible to define given the lack of true negative controls. In sensitivity analyses of SARS-CoV-2 seroprevalence, we excluded samples with very high MFI to the endemic HCoVs, to check if SARS-CoV-2 seroprevalence estimates changed in this sub-set of samples. We defined these samples as those with MFI units to endemic HCoVs that were three or more standard deviations above the MFI to that HCoV antigen in the SARS-CoV-2 negative control samples. The SARS-CoV-2 negative control samples were sera from European adults and likely included some who had a history of exposure to HCoVs but these represented the closest we had to an HCoV naïve population as HCoV infections are thought to be less prevalent in Europe than in Africa[23].

Correlations in IgG responses that could indicate cross-binding were assessed by plotting the MFI units of IgG to each antigen against one-another and calculating the Pearson’s correlation coefficient.

Exploratory analyses examined factors associated with remaining IgG/M seronegative i.e., not seroconverting to SARS-CoV-2 RBD/N-protein during follow-up, using logistic regression, among unvaccinated participants in Goma. There were too few participants who remained seronegative in Masaka and Kambia to perform this analysis there. Additionally, the distributions of log IgG MFI units to HCoVs at baseline were compared among participants who subsequently seroconverted to SARS-CoV-2 during follow up and those who remained negative during follow-up to determine if pre-existing IgG responses to endemic HCoVs predicted risk of acquisition of SARS-CoV-2 infection during the study. Log IgG MFI unit distributions were compared using the Wilcoxon rank-sum test. Linear regression of MFI units was used to determine if there was significant waning of IgG to SARS-CoV-2 RBD/NP over time, controlling for clustering of data by participant.

Among participants who received one or two doses of any COVID-19 vaccine before or during the study, we plotted IgG MFI to SARS-CoV-2 RBD by time since vaccination and by dose, using the recorded date of vaccination in their vaccination records. In an exploratory analysis, we looked at whether seropositivity to SARS-CoV-2 at baseline influenced subsequent post-vaccination SARS-CoV-2 RBD IgG MFI, controlling for the time of sampling since vaccination.

## Results

### The study population

The baseline serosurvey enrolled 196 participants in February 2021 in Goma, DRC, 126 participants in March 2021 in Kambia, Sierra Leone, and 250 participants in July of the same year in Masaka District, Uganda. Retention at month four was 96% in Goma (189/196), 98% in Kambia (123/126) and 92% in Masaka (230/250)(**Figure 1**). The study populations, which included everyone with patient contact at the selected health facilities, were distinct in the different settings (**Table 1**). A high proportion of Sierra Leonean staff had a mid-upper arm circumference (MUAC) of >31cm (79/126; 63%) and known pre-existing conditions (98/126; 78%). At least one contact with a known COVID-19 case over the previous month was reported by 41% (103/250) of Ugandan participants, 27% (52/196) of participants in DRC but only one participant in Sierra Leone.

**Table 1.**
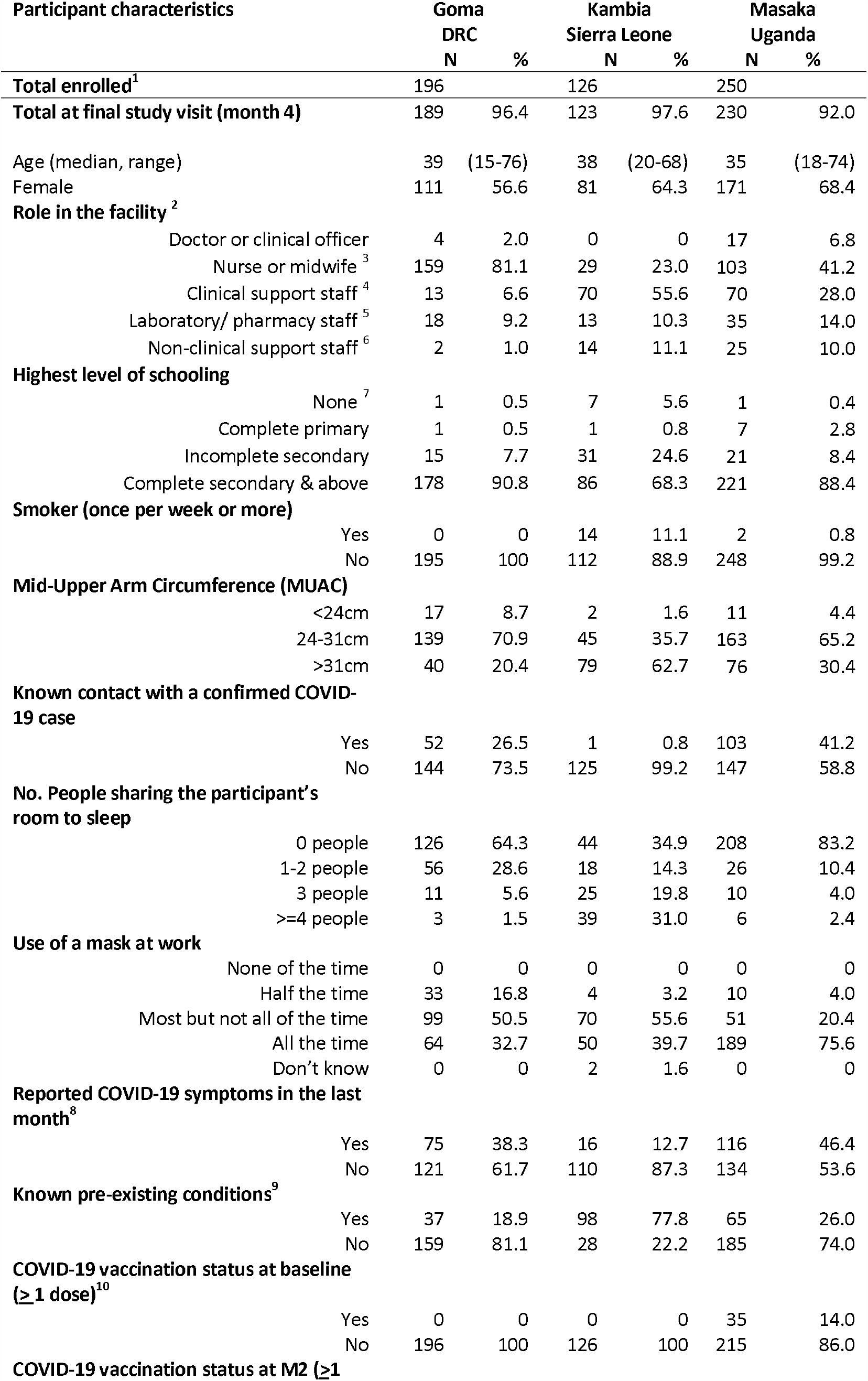
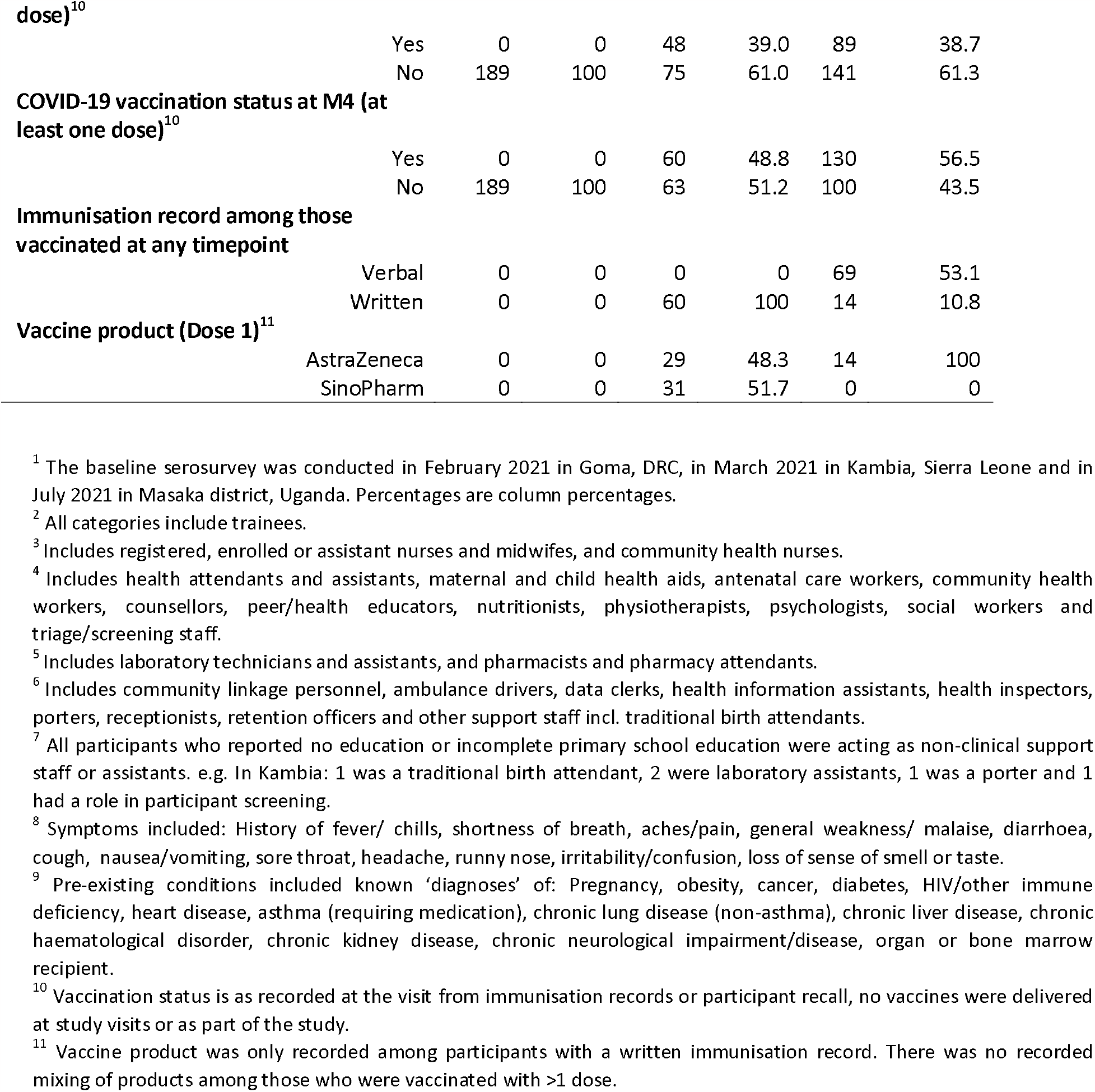
Description of the study population.

**Figure 1.**
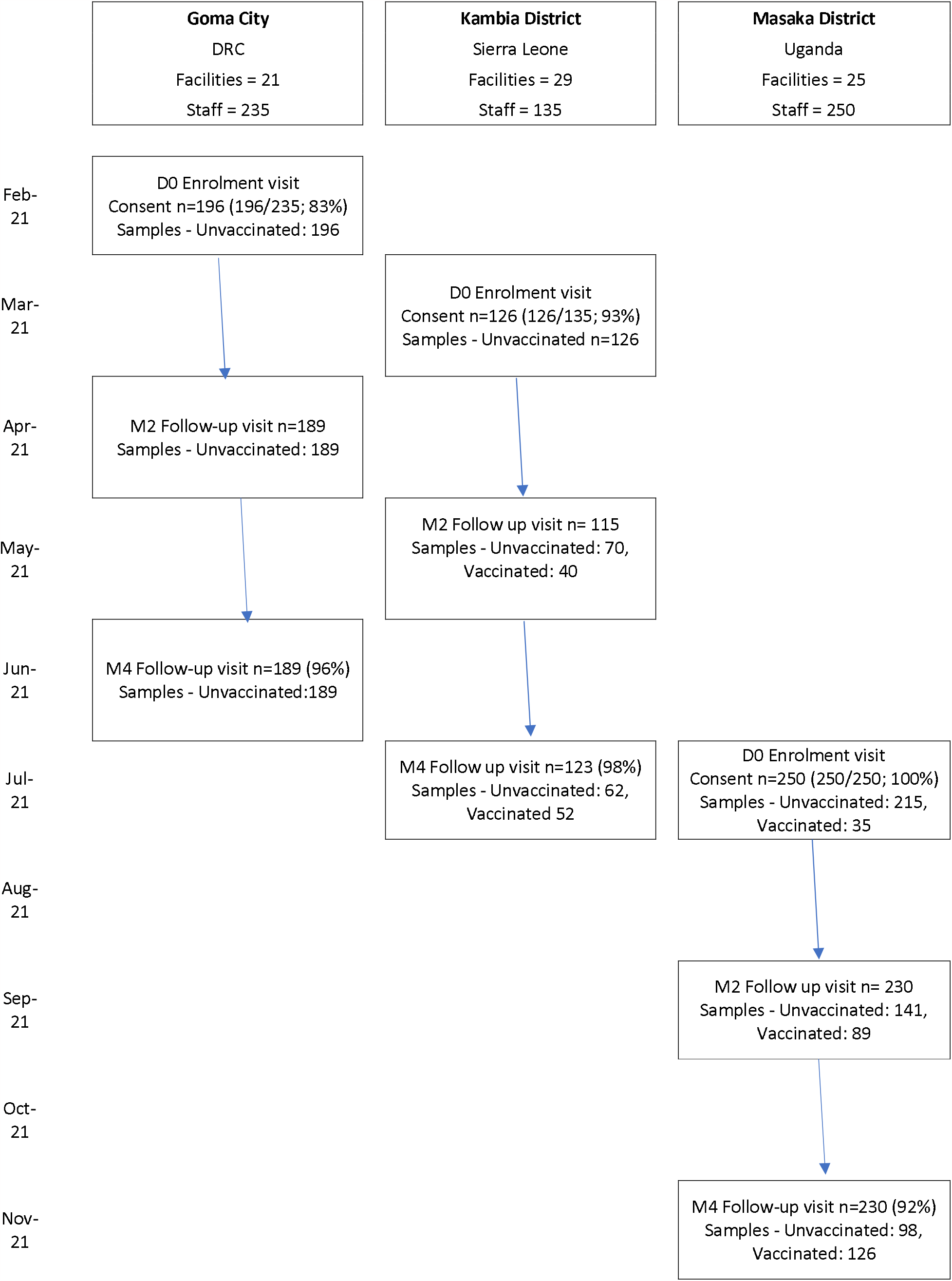
Study timeline and participant flow^1^. ^1^COVID-19 vaccination programmes started in Sierra Leone and Uganda in March 2021, and in DRC in April 2021

**Figure. 1.**
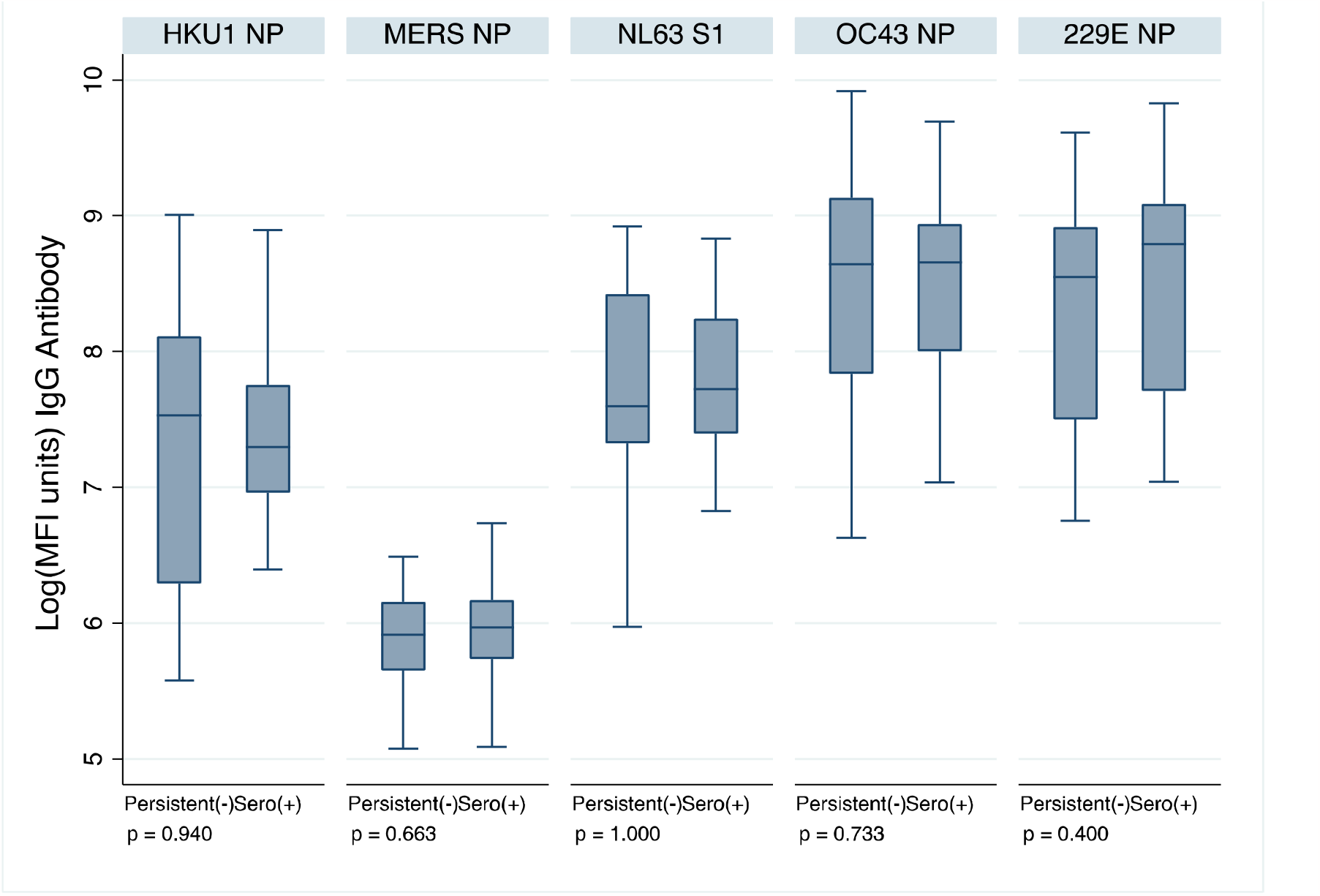
HCoV IgG MFI units at baseline, comparing 23 participants who remained seronegative during follow up with 28 participants who were seronegative at baseline and acquired SARS-CoV-2 infection during follow up (seroconverted to SARS-CoV-2 RBD/NP IgG/M at Month 2 or 4), in Goma. P-values are for a Wilcoxon rank-sum test

The baseline surveys were conducted just prior to the implementation of the national COVID vaccination programmes in in DRC and Sierra Leone; in both settings, vaccinations began in April 2021. In Goma, all study participants remained unvaccinated for the duration of the study; in Kambia, 49% (60/123) of participants had received at least one dose of vaccine by the end of the study (month 4). In Uganda, COVID-19 vaccination began in March 2021; 14% (35/250) of study participants had been vaccinated with at least one dose at the baseline study visit in July and this increased to 56% (130/230) by the final study visit in November 2021.

### SARS-CoV-2 seroprevalence among unvaccinated participants

Among unvaccinated participants, the prevalence of IgG/M to SARS-CoV-2 RBD or N-protein at enrolment was 70% (138/196) in Goma, 89% (112/126) in Kambia and 89% (190/213) in Masaka (Table 2). In Goma, 14% (28/186) of unvaccinated participants with IgG and IgM data at all timepoints, were seronegative at baseline and became seropositive during the four months of the study i.e., seroconverted; 7 (4%) sero-reverted (Suppl. Table 1). In Sierra Leone, 4 of 62 (7%) unvaccinated participants were initially seronegative and seroconverted during follow-up; 9 (15%) were initially seropositive and seroconverted during follow-up (Suppl. Table 1). In Uganda, 4 of 98 (4%) unvaccinated participants were seronegative at baseline and seroconverted during follow up and 3 of 98 (3%) unvaccinated participants were initially seropositive and seroconverted during follow-up (Suppl. Table 1). Among unvaccinated participants, almost all had detectable IgG/IgM to SARS-CoV-2 RBD/NP at least once by the end of the 4 months of follow-up: 88% were seropositive to SARS-CoV-2 at least once in Goma, 100% in Sierra Leone, and 96% in Masaka (**Table 2**, Suppl. Table 1, **Suppl Fig 1**.).

**Table 2.**
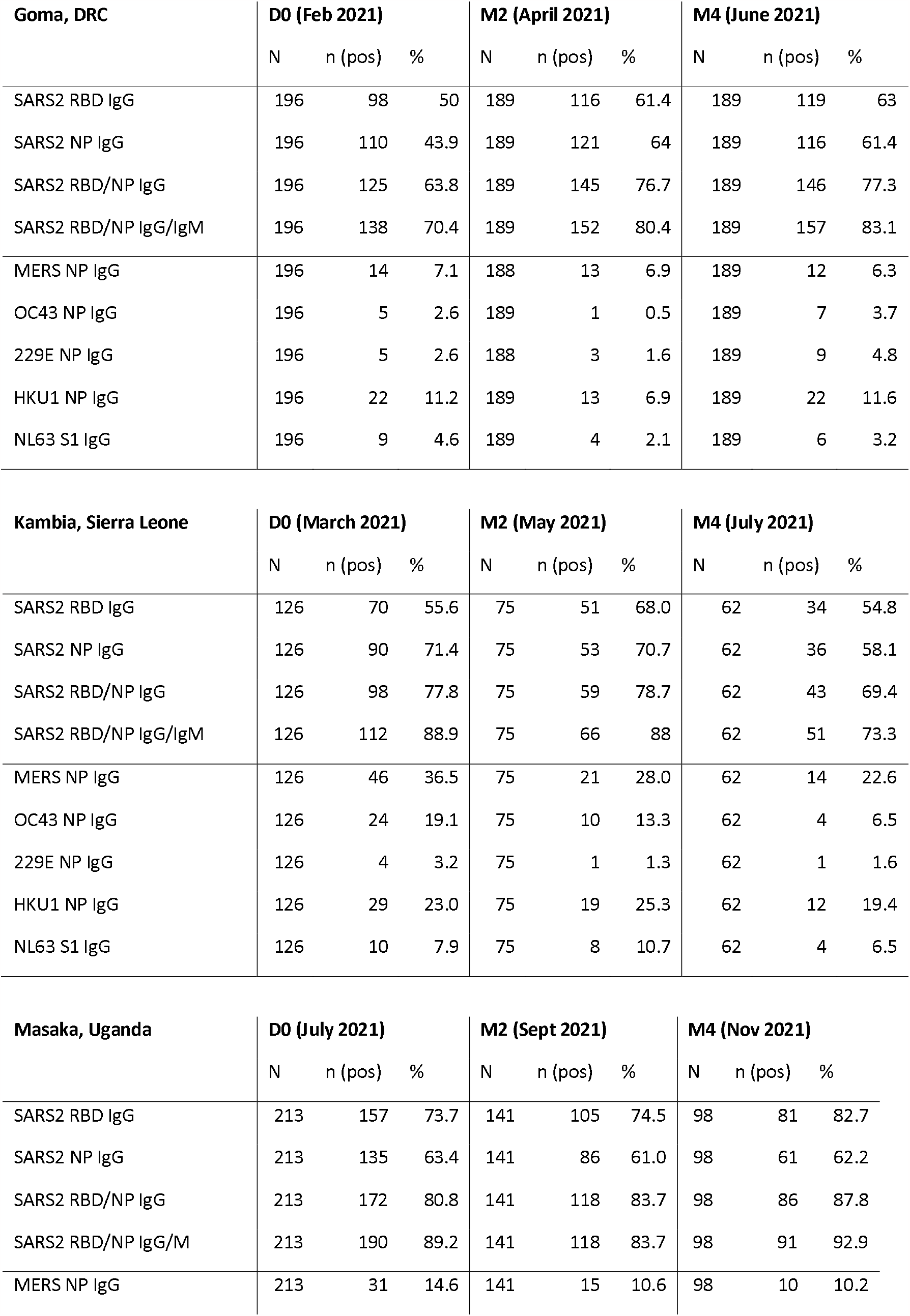
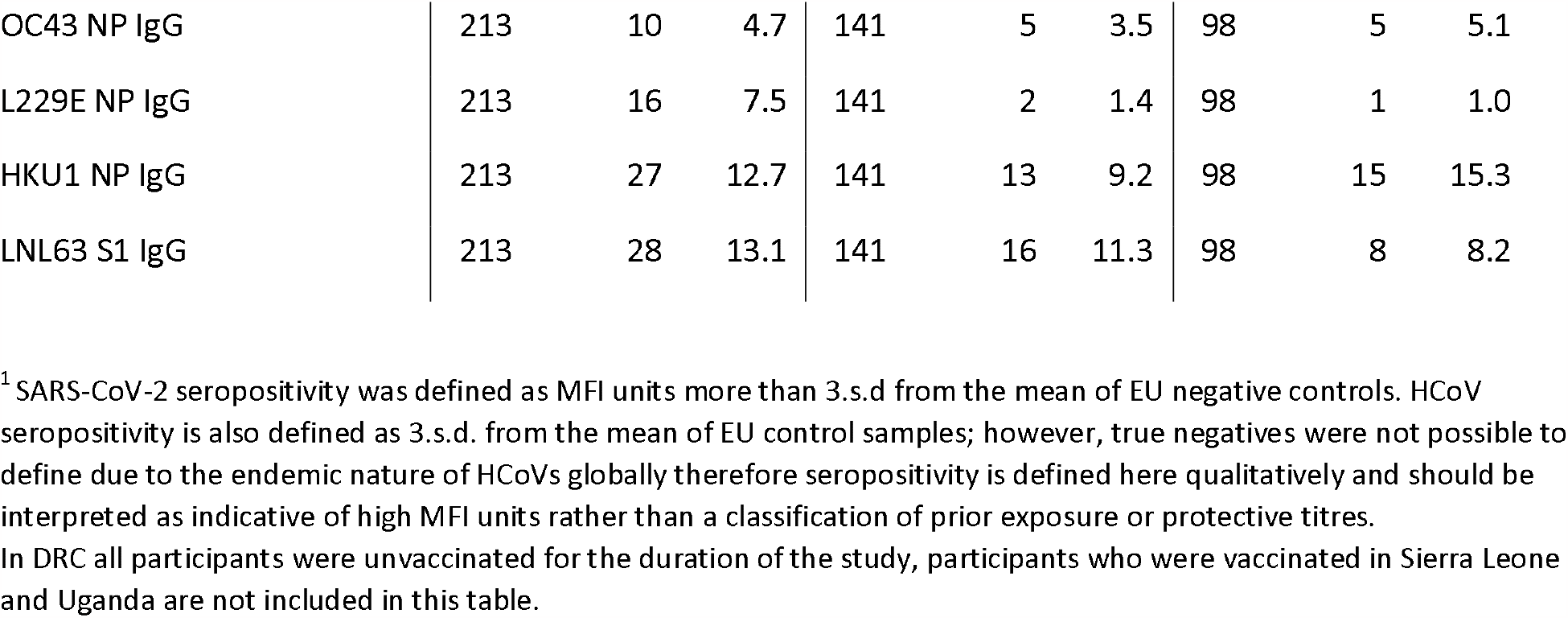
SARS-CoV-2 seroprevalence among unvaccinated participants^1^ and the prevalence of samples with high MFI to other HCoVs.

| **Goma, DRC** | **D0 (Feb 2021)** |  |  | **M2 (April 2021)** |  |  | **M4 (June 2021)** |  |  |
| --- | --- | --- | --- | --- | --- | --- | --- | --- | --- |
|  | N | n (pos) | % | N | n (pos) | % | N | n (pos) | % |
| SARS2 RBD IgG | 196 | 98 | 50 | 189 | 116 | 61.4 | 189 | 119 | 63 |
| SARS2 NP IgG | 196 | 110 | 43.9 | 189 | 121 | 64 | 189 | 116 | 61.4 |
| SARS2 RBD/NP IgG | 196 | 125 | 63.8 | 189 | 145 | 76.7 | 189 | 146 | 77.3 |
| SARS2 RBD/NP IgG/IgM | 196 | 138 | 70.4 | 189 | 152 | 80.4 | 189 | 157 | 83.1 |
| MERS NP IgG | 196 | 14 | 7.1 | 188 | 13 | 6.9 | 189 | 12 | 6.3 |
| OC43 NP IgG | 196 | 5 | 2.6 | 189 | 1 | 0.5 | 189 | 7 | 3.7 |
| 229E NP IgG | 196 | 5 | 2.6 | 188 | 3 | 1.6 | 189 | 9 | 4.8 |
| HKU1 NP IgG | 196 | 22 | 11.2 | 189 | 13 | 6.9 | 189 | 22 | 11.6 |
| NL63 S1 IgG | 196 | 9 | 4.6 | 189 | 4 | 2.1 | 189 | 6 | 3.2 |

| **Kambia, Sierra Leone** | **D0 (March 2021)** |  |  | **M2 (May 2021)** |  |  | **M4 (July 2021)** |  |  |
| --- | --- | --- | --- | --- | --- | --- | --- | --- | --- |
|  | N | n (pos) | % | N | n (pos) | % | N | n (pos) | % |
| SARS2 RBD IgG | 126 | 70 | 55.6 | 75 | 51 | 68.0 | 62 | 34 | 54.8 |
| SARS2 NP IgG | 126 | 90 | 71.4 | 75 | 53 | 70.7 | 62 | 36 | 58.1 |
| SARS2 RBD/NP IgG | 126 | 98 | 77.8 | 75 | 59 | 78.7 | 62 | 43 | 69.4 |
| SARS2 RBD/NP IgG/IgM | 126 | 112 | 88.9 | 75 | 66 | 88 | 62 | 51 | 73.3 |
| MERS NP IgG | 126 | 46 | 36.5 | 75 | 21 | 28.0 | 62 | 14 | 22.6 |
| OC43 NP IgG | 126 | 24 | 19.1 | 75 | 10 | 13.3 | 62 | 4 | 6.5 |
| 229E NP IgG | 126 | 4 | 3.2 | 75 | 1 | 1.3 | 62 | 1 | 1.6 |
| HKU1 NP IgG | 126 | 29 | 23.0 | 75 | 19 | 25.3 | 62 | 12 | 19.4 |
| NL63 S1 IgG | 126 | 10 | 7.9 | 75 | 8 | 10.7 | 62 | 4 | 6.5 |

| **Masaka, Uganda** | **D0 (July 2021)** |  |  | **M2 (Sept 2021)** |  |  | **M4 (Nov 2021)** |  |  |
| --- | --- | --- | --- | --- | --- | --- | --- | --- | --- |
|  | N | n (pos) | % | N | n (pos) | % | N | n (pos) | % |
| SARS2 RBD IgG | 213 | 157 | 73.7 | 141 | 105 | 74.5 | 98 | 81 | 82.7 |
| SARS2 NP IgG | 213 | 135 | 63.4 | 141 | 86 | 61.0 | 98 | 61 | 62.2 |
| SARS2 RBD/NP IgG | 213 | 172 | 80.8 | 141 | 118 | 83.7 | 98 | 86 | 87.8 |
| SARS2 RBD/NP IgG/M | 213 | 190 | 89.2 | 141 | 118 | 83.7 | 98 | 91 | 92.9 |
| MERS NP IgG | 213 | 31 | 14.6 | 141 | 15 | 10.6 | 98 | 10 | 10.2 |
| COVID19 HWI serosurvey manuscript - Tables |  |  |  |  |  |  |  |  |  |
| OC43 NP IgG | 213 | 10 | 4.7 | 141 | 5 | 3.5 | 98 | 5 | 5.1 |
| L229E NP IgG | 213 | 16 | 7.5 | 141 | 2 | 1.4 | 98 | 1 | 1.0 |
| HKU1 NP IgG | 213 | 27 | 12.7 | 141 | 13 | 9.2 | 98 | 15 | 15.3 |
| LNL63 S1 IgG | 213 | 28 | 13.1 | 141 | 16 | 11.3 | 98 | 8 | 8.2 |

<sup>1</sup> SARS-CoV-2 seropositivity was defined as MFI units more than 3.s.d from the mean of EU negative controls. HCoV seropositivity is also defined as 3.s.d. from the mean of EU control samples; however, true negatives were not possible to define due to the endemic nature of HCoVs globally therefore seropositivity is defined here qualitatively and should be interpreted as indicative of high MFI units rather than a classification of prior exposure or protective titres.
In DRC all participants were unvaccinated for the duration of the study, participants who were vaccinated in Sierra Leone and Uganda are not included in this table.

### Correlations between IgG responses to SARS-CoV-2 and other HCoVs

Across all three settings, several samples demonstrated high MFI to MERS NP, i.e., responses at least 3 standard deviations higher than negative controls. Serum IgG bound to MERS NP, despite no known circulation of MERS or clinical presentation of the disease in these settings. Additionally, the level of binding differed by site: In Goma, 7% of samples contained IgG that bound to MERS NP, in Uganda 14% and in Sierra Leone 37% (Table 2). There were no statistically significant correlations between IgG MFI units to MERS-NP and SARS-CoV-2 NP. To determine why the prevalence of IgG binding to MERS-NP differed by site we looked at correlations between IgG MFI to MERS NP protein and other HCoVs. In Goma, IgG MFI units to MERS-NP correlated with OC43-NP and 229E-NP. In Uganda, MFI to MERS-NP correlated with OC43-NP, HKU1-NP, and NL63 S1. In Sierra Leone, where MERS-NP binding was most prevalent, but the sample size was smallest, no correlations were seen between MFI units to MERS NP protein and other HCoVs (**suppl. Fig. 2, suppl. Table 2**).

Between 1 and 19% of samples across sites and timepoints exhibited high IgG MFI units (at least 3.s.d. higher than the MFI units in SARS-CoV-2 negative control samples) to endemic coronaviruses (Table 2). At the baseline visit, there were some correlations between MFI units of IgG to SARS-CoV-2 NP and MFI units of IgG to NP of the endemic HCoVs. In samples from Goma, MFI units of IgG to SARS-CoV-2 NP correlated with OC43-NP and 229E-NP, and IgG to SARS-CoV-2 RBD correlated with IgG to NL63 S1-protein. In samples from Uganda, MFI units of IgG to SARS-CoV-2 NP correlated with IgG MFI units to 229E-NP only. In Sierra Leone no significant correlations were detected (**Suppl. Fig. 3, Suppl. Table 3**).

Due to this indication of cross-binding of IgG we conducted a sensitivity analysis, excluding samples with high MFI to each HCoV in turn, and there were no significant differences in the estimates of SARS-CoV-2 seroprevalence estimated across the whole sample set and those estimated in a restricted dataset of samples with lower MFI to each HCoV (**suppl. Table 4**).

### Pre-existing HCoV IgG and ‘acquisition’ of SARS-CoV-2 in unvaccinated participants

In Goma, 28 of 186 (14%) unvaccinated participants with samples at every timepoint, were initially seronegative but seroconverted or ‘acquired’ SARS-CoV-2 IgG during the study; 23 (12%) participants remained seronegative throughout follow-up. There were no significant differences in IgG MFI units to endemic HCoVs at baseline in participants who subsequently went on to acquire SARS-CoV-2 IgG/M compared to those who remained seronegative throughout follow-up (**Fig. 2**). Too few participants remained seronegative during follow up in Sierra Leone and Uganda to contribute to this analysis.

**Fig 2.**
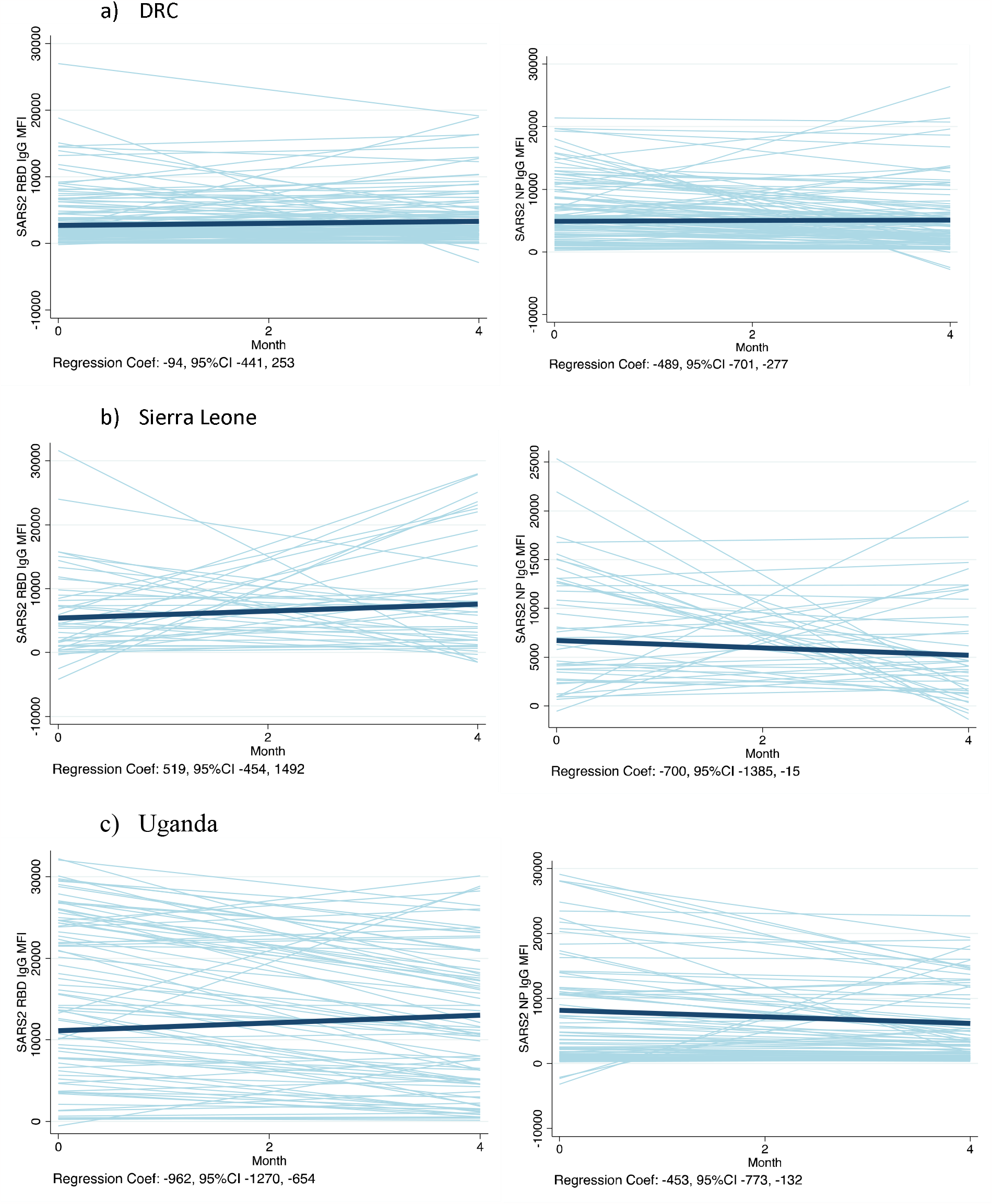
Trends in IgG MFI units to SARS-CoV-2 RBD and NP over time, among those persistently positive for IgG or IgM to RBD or NP^1^. ^1^ Spaghetti plots represent the predicted linear trends for each participant in light blue lines, and the bold line indicates the average slope across participants. The actual linear regression coefficient (slope), controlling for clustering of data by participant with random effects and robust standard errors, is indicated below each graph with its 95%CI. The number of participants included are as follows: DRC n=119; Sierra Leone n=41; Uganda n=83.

### Rate of MFI unit decay among participants unvaccinated and seropositive at baseline

In an analysis of the trend in IgG MFI units over time among unvaccinated participants who were persistently seropositive, linear regression coefficients indicated MFI waning in Uganda, but not in the DRC, nor in Sierra Leone (**Figure 3**), controlling for clustering of data by participant with random effects and robust standard errors. In contrast, IgG to NP waned over time in all three settings (**Figure 3**).

**Fig 3.**
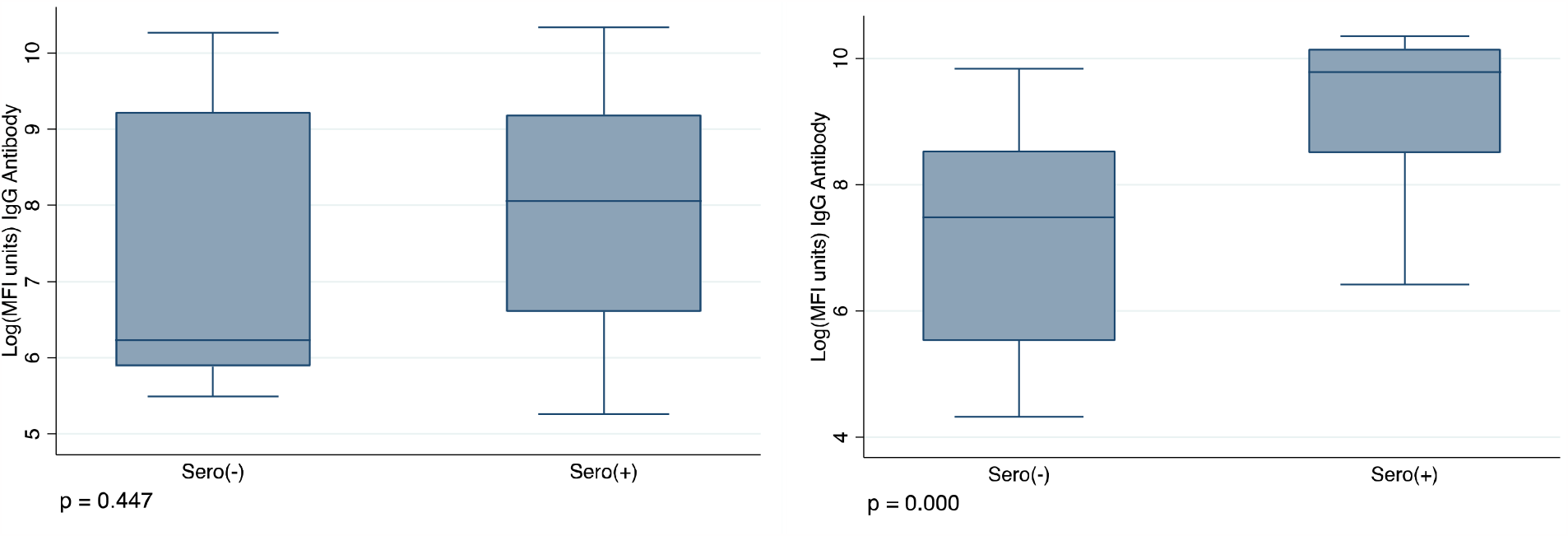
IgG to SARS-CoV-2 RBD >14 days post-vaccination with 1 dose, by serostatus at baseline. P-values from the Wilcoxon Rank Sum test. ^1^ In Sierra leone just 5 seronegative participants at baseline (i.e. seronegative for IgM and IgG to RBD and NP) had a recorded vaccination date and a blood sample >14 days after vaccination (Sero(-) group n=4), 44 participants were seropositive at baseline (i.e. were seropositive for IgM or IgG to RBD or NP) had a recorded vaccination date and a blood sample >14 days after vaccination (Sero(+) group n=44); median of 47 days between vaccination and a study sample (IQR 28-56, range 15-79). ^2^ In Uganda, just 12 unvaccinated and seronegative participants at baseline had a recorded vaccination date after baseline and a blood sample >14 days after vaccination, 96 participants were seropositive at baseline and had a recorded vaccination date after baseline, with a blood sample >14 days after vaccination. Median interval between vaccination and blood sample was 42 days (IQR 28-88).

### Factors associated with remaining seronegative to SARS-CoV2 among unvaccinated

In univariable analyses, gender was the only characteristic that was associated with remaining seronegative when compared to participants with at least one seropositive sample in Goma. Male participants had 5.9 times higher odds (95%CI 2-16) of remaining seronegative compared to female participants (**Suppl. Table 5**). In this population, there were no differences in levels of IgG to other endemic HCoVs at baseline among those who remained persistently negative and those who seroconverted (**Suppl. Fig 4**). Too few participants remained seronegative by the end of the study in Sierra Leone and Uganda to complete this analysis in those datasets.

### Serological responses to vaccination

In Sierra Leone, 37 of 40 (93%) participants who were vaccinated between baseline and month 2 surveys were seropositive for IgG/M to RBD/N protein by month 2, but 4 of these participants had sero-reverted by month 4. Eight of the 12 (67%) participants vaccinated between month 2 and 4 had seroconverted by month 4 (**Suppl. Table 6**). When analysing

IgG responses to SARS-CoV-2 RBD alone, by time since first vaccination, 19/ 78 (24%) of IgG responses to RBD were below the threshold defined as ‘seropositive’. The median time since vaccination was 55 days (range 20-93 days) for these samples (**Suppl. Fig. 5**). After dose 2, 11/33 participants (33%) had not mounted an IgG response to RBD that would be defined as ‘seropositive’ in this study. These samples were taken a median of 36 days post-vaccination (range 10-76).

In Uganda, among participants vaccinated before the baseline survey in Uganda, all 34 (100%) were still seropositive by month 4 of follow up. Among those vaccinated between the baseline and month 2 survey, 98% were seropositive by month 4 and among those vaccinated between month 2 and month 4 surveys, 88% were seropositive at month 4 (**Suppl. Table 6)**. When analysing IgG responses to SARS-CoV-2 RBD, by time since vaccination with the first dose, 19/128 (15%) of IgG responses to RBD were below the threshold defined as ‘seropositive’ – median time since vaccination was 26 days (IQR 14-87) for these samples (**Suppl. Fig. 5**).

None of the participants in Goma reported receiving vaccination by the end of the study.

### Prevalence of hybrid immunity and its influence on vaccine responses

In Sierra Leone, 70% (43/60) of the participants who were vaccinated during the study follow-up period, had evidence of natural exposure at baseline, prior to vaccination, and therefore were likely to have hybrid immunity. There was no evidence of a difference in MFI units after dose 1, comparing those participants who were seronegative at baseline with those seropositive at baseline, restricting to only samples taken at least 14 days after vaccination and controlling for the timing of sample collection since vaccination, but the sample size was small (linear reg coefficient −350; 95%CI −7388 – 6686; **Fig. 4**).

In Uganda, 75% (96/120) of the participants who were unvaccinated at baseline and were vaccinated during the study follow-up period, had evidence of natural exposure at baseline and are therefore likely to have hybrid immunity. There was evidence of significantly higher IgG MFI after dose 1 among those who were seropositive at baseline compared to those who were seronegative at baseline, restricting to only samples taken at least 14 days after vaccination and controlling for the timing of sample collection since vaccination (linear reg coefficient 11904, 95%CI 6233-17576; **Fig 4**).

## Discussion

We documented 70-90% seroprevalence to SARS-CoV-2 in February-July 2021 among unvaccinated staff working at health facilities in three distinct settings in sub-Saharan Africa. Very few of the participants who were seronegative at the beginning of the study remained seronegative 4 months later, indicating a high force of infection. Literature indicates that SARS-CoV-2 and other HCoVs are homologous enough for their corresponding antibodies to exhibit cross-binding on ELISA/ multiplex assays[7, 9, 12] and that this cross-binding can be neutralising[24]. We found evidence that IgG to SARS-CoV-2 N-protein may cross-bind N-proteins of other endemic HCoVs and that IgG to endemic HCoVs may cross-bind MERS N-protein. It is impossible to determine whether binding IgG results from genuine prior exposure to endemic HCoVs or cross-reactivity within the assay. To attempt to control for this we conducted a sensitivity analysis of SARS-CoV-2 seroprevalence, stratified by HCoV sero-status and found no difference in our estimates of SARS-CoV-2 seroprevalence. However, numbers of participants with high MFI to endemic HCoVs in some of the comparisons are small.

We found evidence of waning of natural IgG to SARS-CoV-2 RBD in Uganda but not in DRC or Sierra Leone, perhaps the lack of waning was due to natural boosting with repeated re-exposure, a limitation of the study was that we did not collect swab samples for virological analyses. Cross-sectional seroprevalence estimates only provide a snapshot of a dynamic epidemic. Longitudinal follow-up documents how participants fluctuate between seropositive and seronegative status over time. Pre-existing HCoV IgG did not predict the likelihood of sero-acquisition of SARS-CoV-2 during follow-up in unvaccinated participants, perhaps because of the high force of infection in the study settings. None of the participants reported symptoms during follow-up and so we could not analyse the likelihood of symptomatic infection by HCoV serostatus at baseline. Prior studies have indicated prior exposure to HCoVs may be protective against symptomatic SARS-CoV-2[13, 14].

A number of participants in Sierra Leone were seronegative for IgG to RBD three or more weeks post-vaccination, this wasn’t as much of a problem in Uganda. It may be that participants required more time post-vaccination to mount an IgG response to vaccination (given the observational nature of this study we had no control over when they received vaccination in relation to study visits), or IgG to RBD alone may not be a good marker of vaccination status in our settings. Three-quarters (70-75%) of those vaccinated during follow-up in Sierra Leone and Uganda had evidence of natural immunity prior to vaccination and therefore had hybrid immunity post-vaccination. Prior exposure to SARS-CoV-2 correlated with better post-vaccination IgG to RBD in Uganda but not in Sierra Leone, numbers were small in this group and the analysis was limited by the lower IgG responses to vaccination in Sierra Leone. The prevalence of hybrid immunity should be built into predictions of the severity of future waves of the pandemic in these settings[2].

This was an observational cohort, the high SARS-CoV-2 seroprevalence at baseline and the start of the COVID-19 vaccination programme during follow up resulted in several analyses containing small numbers of participants and therefore results should be interpreted with caution. Given the number of antigens, immunoglobulins and timepoints, some of the apparent significant findings could have arisen by chance. We had no SARS-CoV-2 virological data and therefore cannot validate our definitions of ‘acquisition of infection’ with antigen data. There may be evidence of more natural boosting (re-exposure) in Sierra Leone and Goma as there was no evidence of waning of MFI units over time among seropositive participants who remained unvaccinated, however, without virological data and/or an estimate of the time of infection, it was difficult to make any conclusions here. SARS-CoV-2 transmission was heterogeneous across time and geography, the findings here do not necessarily represent the overall situation among health care staff across the DRC, Uganda and Sierra Leone.

## Conclusion

There was substantial transmission of SARS-CoV-2 among staff at health care facilities in Goma DRC, Masaka District, Uganda, and Kambia District, Sierra Leone over a four-month period. This will have conferred some natural immunity to infection and subsequent vaccination will confer hybrid immunity. The prevalence of natural infection could be used to inform vaccination policy, and prediction models of the impact of subsequent waves of infection in these settings.

## Data Availability

All data produced in the present study are available upon reasonable request to the authors

## Conflict of Interest

Authors declare no conflicts of interest

## Funding

This project was funded by a UKRI (MRC), DHSC (NIHR) research grant (GEC1017,

MR/V029363/1, PI Katherine Gallagher). The funder was not involved in study design; in the collection, analysis, and interpretation of data; in the writing of the report; and in the decision to submit the article for publication.

## Ethical Approval Statement

This study was approved by the Comite National d’Ethique de la Sante (CNES) of the DRC, the Sierra Leone Ethics and Scientific Review committee, the Uganda Virus Research Institute Research Ethics Committee, the Uganda National Council for Science and Technology, the London School of Hygiene and Tropical Medicine Ethics Committee and local health authorities in each area. Written informed consent was obtained from volunteers prior to participation.

## Acknowledgements

We thank the data collection teams, the health facility staff and the Ministry of Health officials in the study countries for facilitating the collection of this data. We thank Public Health England and NIBSS for supplying control samples.

## Notes

### Competing Interest Statement

The authors have declared no competing interest.

### Author Declarations

This study was approved by the Comite National d Ethique de la Sante (CNES) of the DRC, the Sierra Leone Ethics and Scientific Review committee, the Uganda Virus Research Institute Research Ethics Committee, the Uganda National Council for Science and Technology, the London School of Hygiene and Tropical Medicine Ethics Committee and local health authorities in each area. Written informed consent was obtained from volunteers prior to participation.

