## Supplementary material for "Prevalence of IgG and IgM to SARS-CoV-2 and other human coronaviruses in The Democratic Republic of Congo, Sierra Leone and Uganda: A Longitudinal Study": Suppl.

### Supplementary information

Suppl. Figure. 4. HCoV IgG MFI units at baseline, comparing those who were seropositive to SARS-CoV-2 at least once during the study (‘0’ below, n=163) with those remained persistently seronegative to SARS-CoV-2 IgG/M to RBD/ N (‘1’ below, n=23), in Goma. ..17

Suppl. Figure 5. Trends in IgG MFI to SARS-COV-2 RBD by time since vaccination in Sierra Leone and Uganda. ..........................................................................................19

Suppl. Table 7: Analyses and sample sizes ...........................................................................20

**Suppl. Table 1. Serological profiles of unvaccinated participants over time**

| Luminex IgG/IgM RBD/NP profile | M0 | M2 | M4 | DRC (N=186) | | SL (N=62) | | UG (N=98) | |
|---|---|---|---|---|---|---|---|---|---|
| | | | | n | % | n | % | n | % |
| 1 – Persistently positive | positive | positive | positive | 119 | 64.0 | 41 | 66.1 | 83 | 84.7 |
| 2 - Seroreversion | positive | positive | negative | 5 | 2.7 | 8 | 12.9 | 2 | 2.0 |
| 3 - Seroreversion | positive | negative | negative | 2 | 1.1 | 1 | 1.6 | 1 | 1.0 |
| 4 – Reinfection[1] | positive | negative | positive | 8 | 4.3 | 6 | 9.7 | 4 | 4.1 |
| 5 – Persistently negative | negative | negative | negative | 23 | 12.4 | 0 | 0 | 4 | 4.1 |
| 6 - Acquisition | negative | negative | positive | 4 | 2.2 | 0 | 0 | 3 | 3.1 |
| 7 - Acquisition | negative | positive | positive | 24 | 12.9 | 4 | 6.5 | 1 | 1.0 |
| 8 – Acquisition[1] | negative | positive | negative | 1 | 0.0 | 2 | 3.2 | 0 | 0.0 |

[1]This could also represent misclassification of the M2 sample. Participants included in this table had to have available IgG and IgM data for RBD and NP for every timepoint and had to remain unvaccinated for the duration of the study.

**Suppl. Figure 1. Reverse Cumulative distribution curves – MFI of IgG to SARS-CoV-2 receptor binding domain (RBD) and nucleocapsid protein (NP) – among unvaccinated**

i. DRC

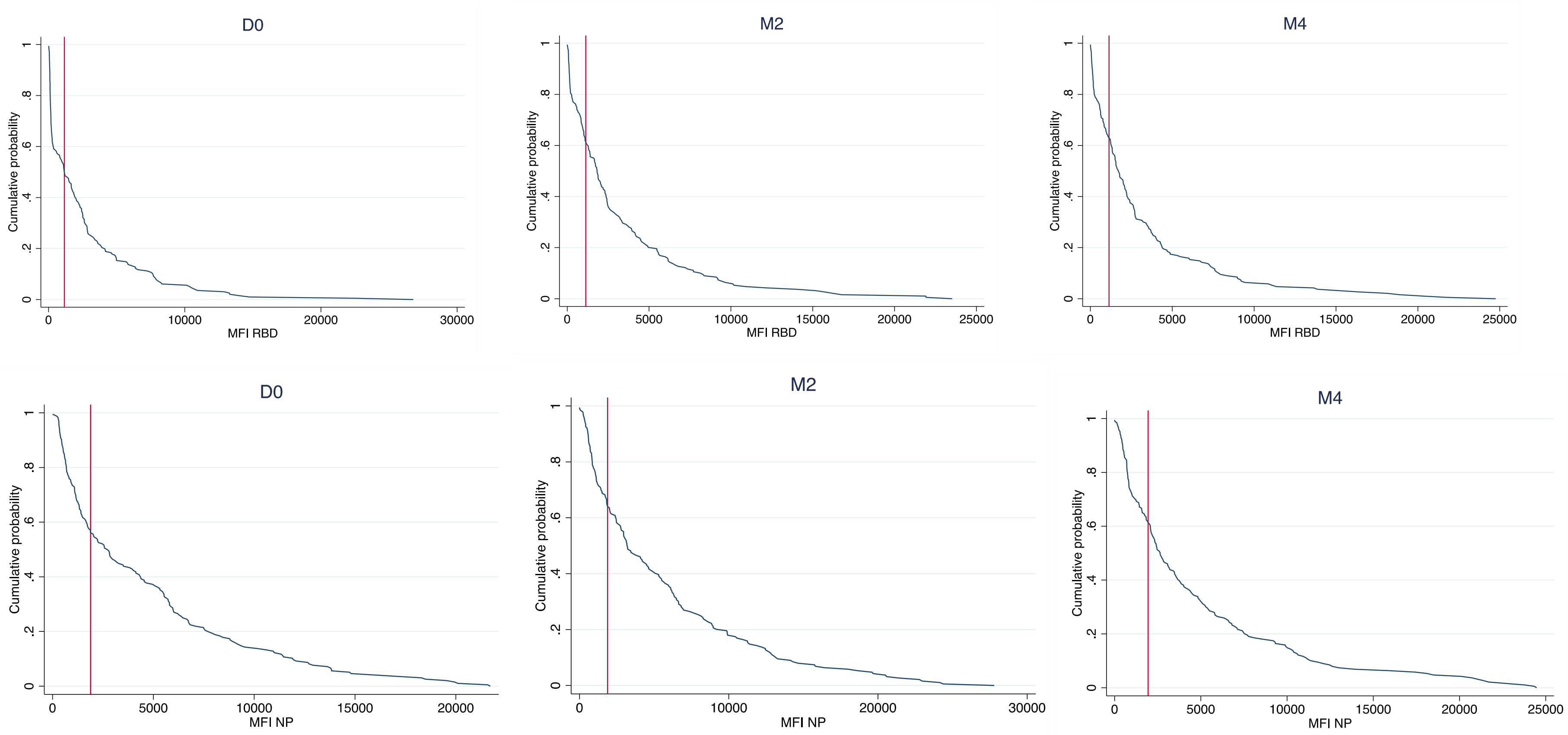

ii. Sierra Leone

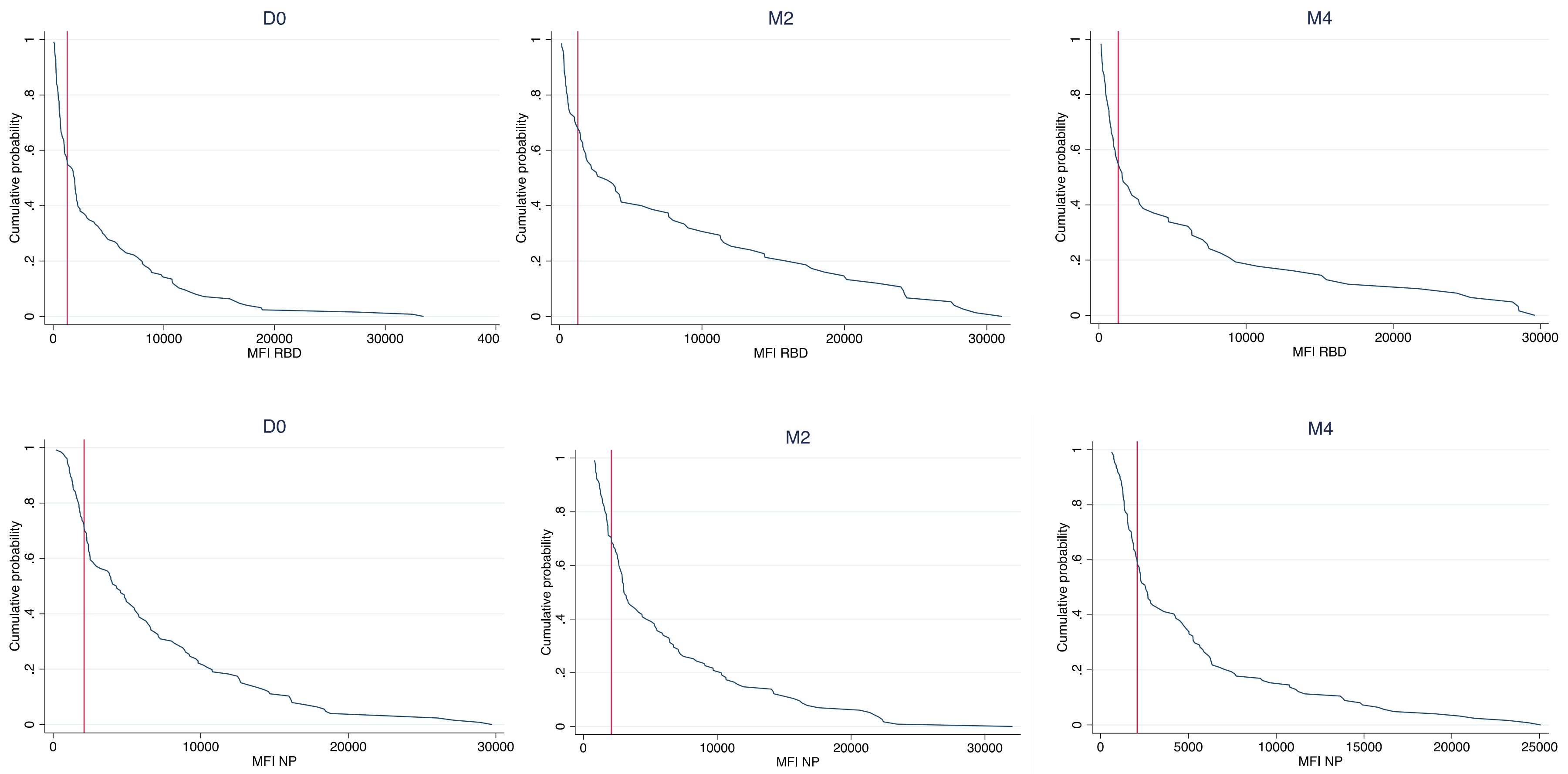

iii. Uganda

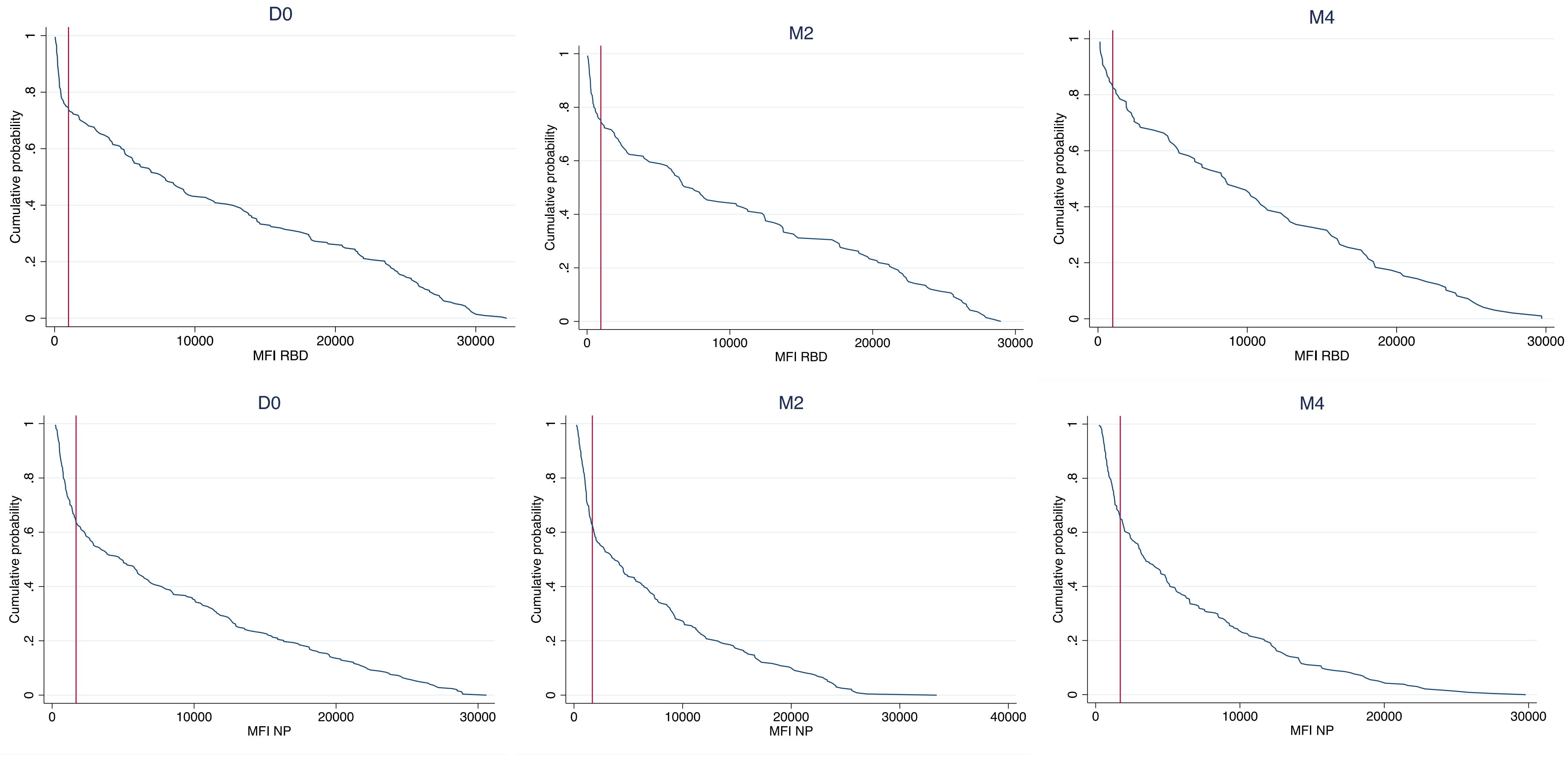
D0
M2
M4
Cumulative probability
MFI RBD
D0
M2
M4
Cumulative probability
MFI NP

**Suppl. Figure 2. Correlation between MERS NP and other HCoVs at baseline. Line: line of best fit**

a) DRC *excluding 1 value of MERS NP MFI > 10,000

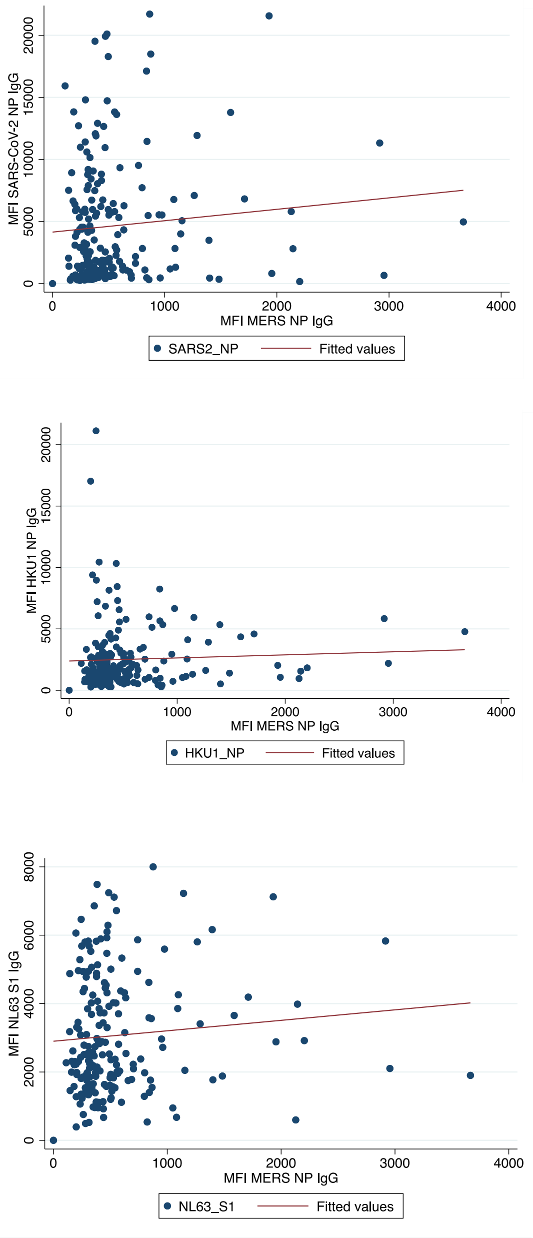

b) Sierra Leone

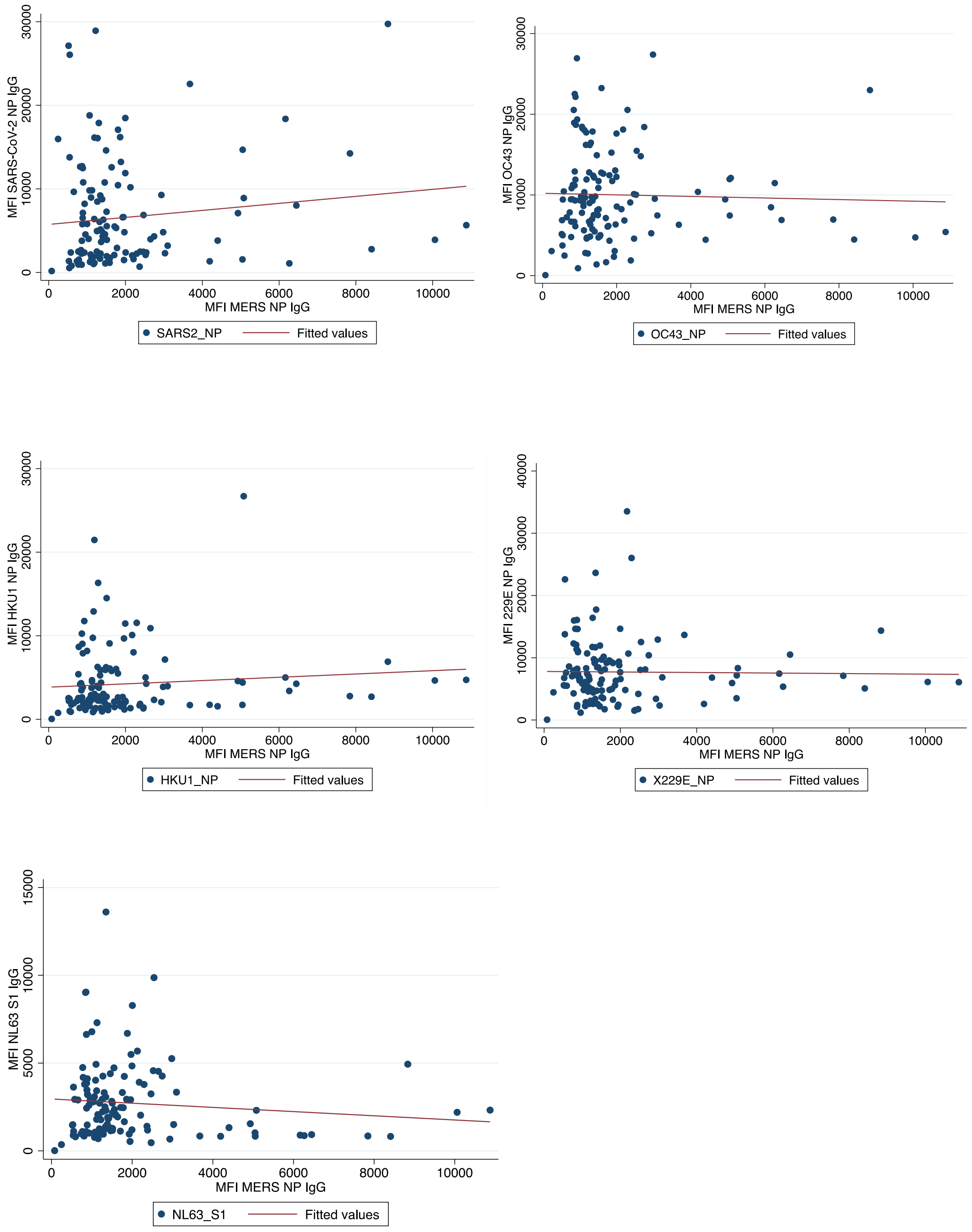

c) Uganda

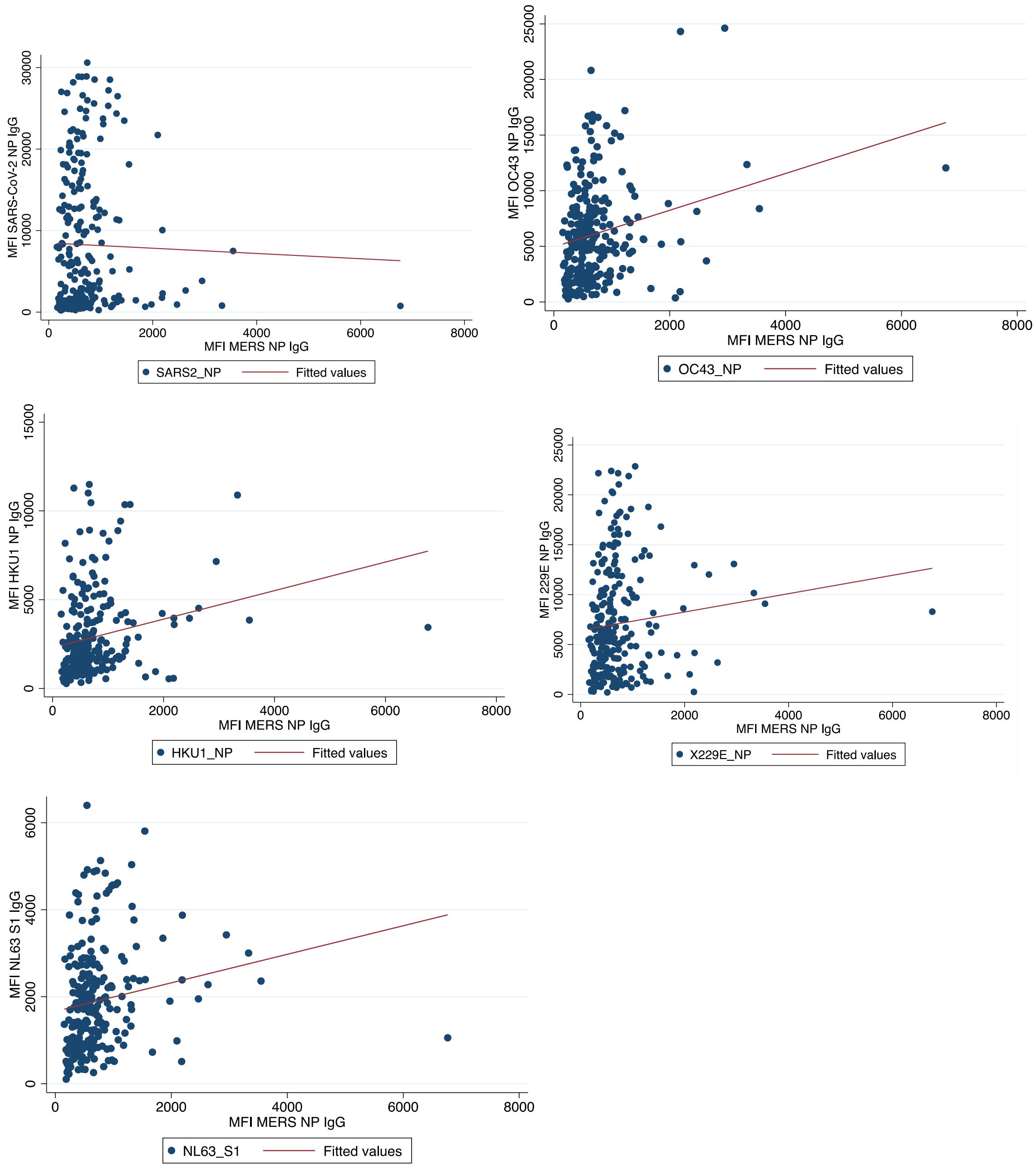

MFI SARS-CoV-2 NP IgG
0
10000
20000
30000
0
2000
4000
6000
8000
MFI MERS NP IgG
SARS2_NP
Fitted values
MFI OC43 NP IgG
0
5000
10000
15000
20000
25000
0
2000
4000
6000
8000
MFI MERS NP IgG
OC43_NP
Fitted values
MFI HKU1 NP IgG
0
5000
10000
15000
0
2000
4000
6000
8000
MFI MERS NP IgG
HKU1_NP
Fitted values
MFI 229E NP IgG
0
5000
10000
15000
20000
25000
0
2000
4000
6000
8000
MFI MERS NP IgG
X229E_NP
Fitted values
MFI NL63 S1 IgG
0
2000
4000
6000
0
2000
4000
6000
8000
MFI MERS NP IgG
NL63_S1
Fitted values

**Suppl. Table 2. Correlation coefficients of MFI units of IgG to MERS-NP and HCoVs among all samples, and MERS-NP with SARS2 among the unvaccinated**

| Antigen | Corr coef. IgG to MERS NP MFI | p-value | n |
|---|---|---|---|
| **DRC** | | | |
| SARS-CoV-2 NP | 0.093 | 0.1959 | 195 |
| OC43 NP | 0.148 | 0.0389 | 195 |
| HKU1 NP | 0.048 | 0.5088 | 195 |
| X229E NP | 0.158 | 0.0275 | 195 |
| NL63 RBD | 0.089 | 0.2161 | 195 |
| **Sierra Leone** | | | |
| SARS-CoV-2 NP | 0.098 | 0.2751 | 126 |
| OC43 NP | -0.033 | 0.7122 | 126 |
| HKU1 NP | 0.0931 | 0.3 | 126 |
| X229E NP | -0.016 | 0.8589 | 126 |
| NL63 RBD | -0.106 | 0.239 | 126 |
| **Uganda** | | | |
| SARS-CoV-2 NP | -0.046 | 0.503 | 213 |
| OC43 NP | 0.239 | 0.001 | 248 |
| HKU1 NP | 0.2141 | 0.0007 | 248 |
| X229E | 0.107 | 0.0944 | 248 |
| NL63 RBD | 0.1720 | 0.007 | 248 |

**Suppl. Figure 3. Correlations between MFI of IgG to SARS-CoV-2 NP/RBD and MFI of IgG to HCoVs among the unvaccinated at enrolment**

a. DRC

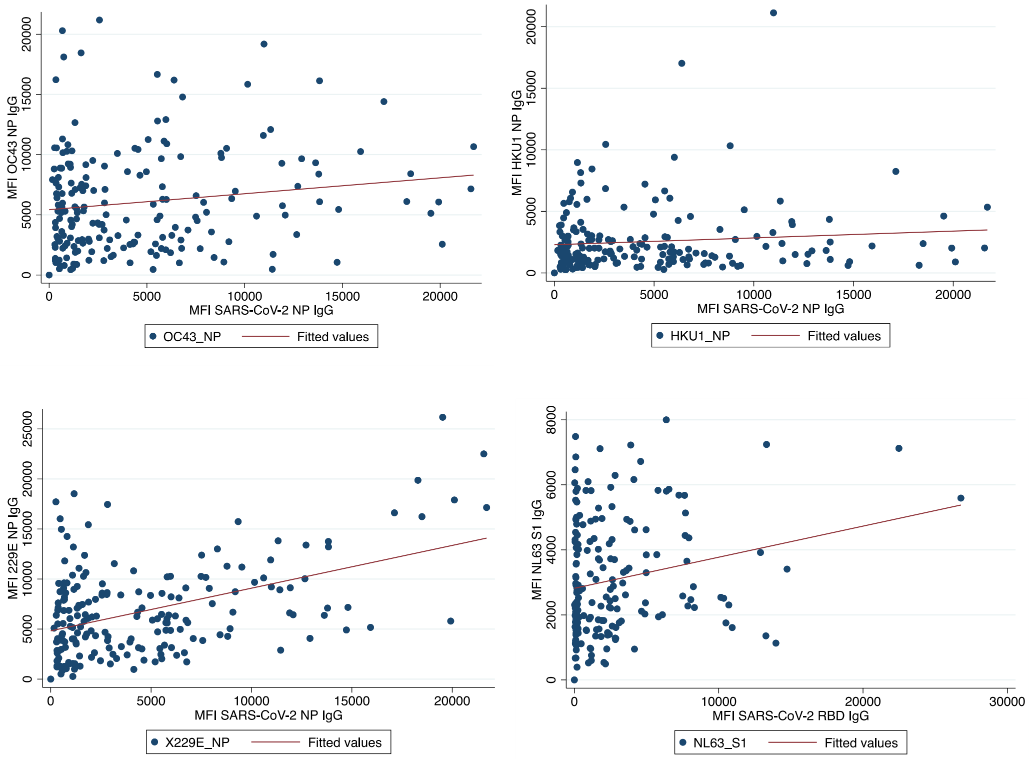

b. Sierra Leone

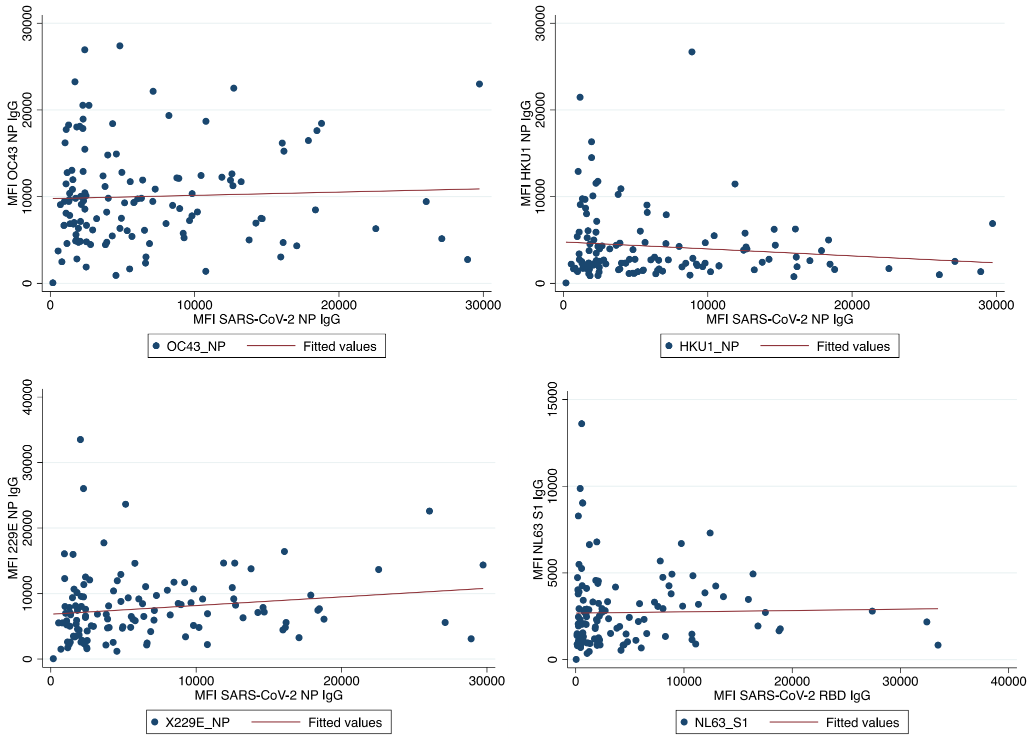

c.  Uganda

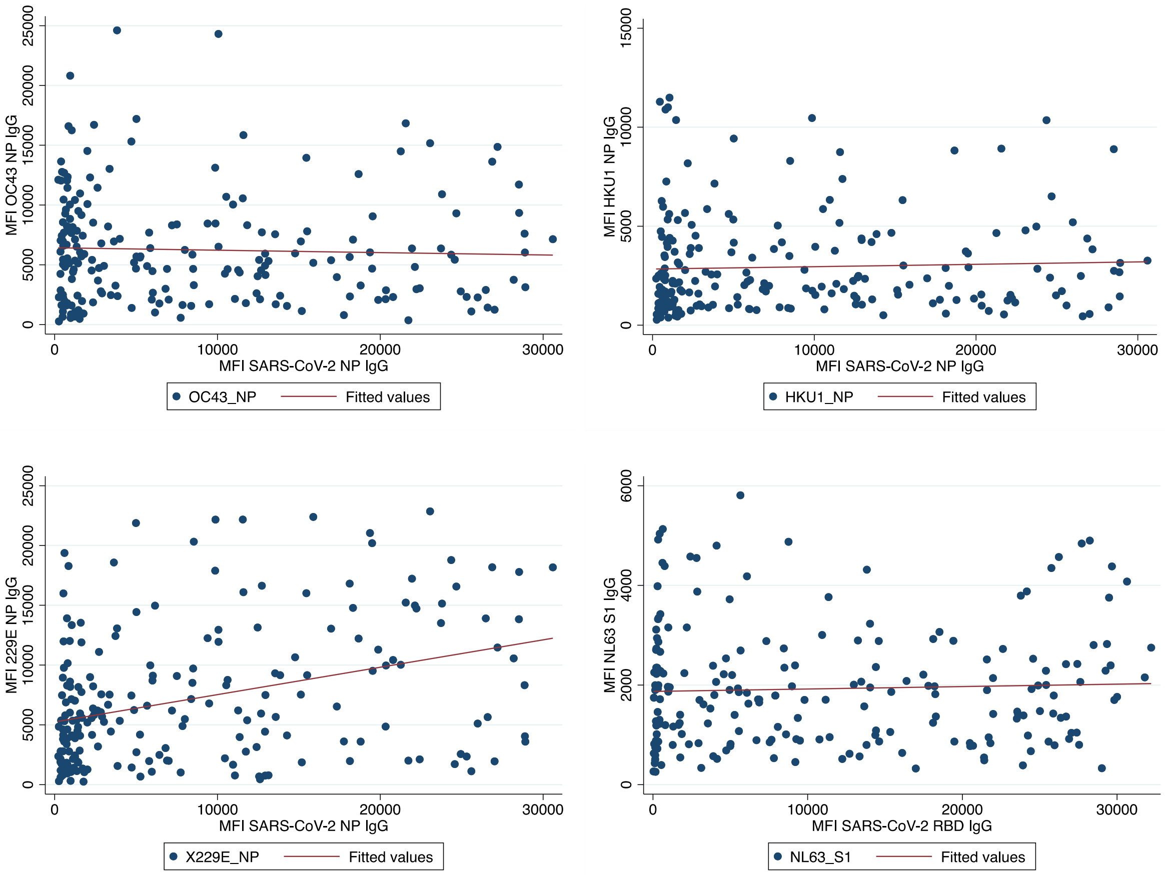
MFI OC43 NP IgG
MFI SARS-CoV-2 NP IgG
OC43_NP
Fitted values
MFI HKU1 NP IgG
MFI SARS-CoV-2 NP IgG
HKU1_NP
Fitted values
MFI 229E NP IgG
MFI SARS-CoV-2 NP IgG
X229E_NP
Fitted values
MFI NL63 S1 IgG
MFI SARS-CoV-2 RBD IgG
NL63_S1
Fitted values

**Suppl. Table 3. Correlation coefficients of MFI units to SARS-CoV-2 NP / RBD protein and HCoVs, among the unvaccinated, at the time of enrolment**

| Antigen | HCoV 'seroprevalence' D0 among unvaccinated | Corr coef. IgG to SARS-COV-2 NP MFI | p-value | n |
|---|---|---|---|---|
| **DRC** | | | | |
| OC43-NP | 2.6 | 0.1513 | 0.0342 | 196 |
| X229E- NP | 2.6 | 0.4608 | <0.001 | 196 |
| HKU1- NP | 11.2 | 0.1024 | 0.1534 | 196 |
| NL63 – S1* | 4.6 | 0.2120 | 0.0029 | 196 |
| MERS-NP | 7.14 | 0.093 | 0.1959 | 196 |
| **Sierra Leone** | | | | |
| OC43-NP | 19.1 | 0.0426 | 0.6358 | 126 |
| X229E- NP | 3.2 | 0.1654 | 0.0642 | 126 |
| HKU1- NP | 23.0 | -0.127 | 0.1558 | 126 |
| NL63 – S1* | 7.9 | 0.0214 | 0.8120 | 126 |
| MERS-NP | 36.5 | 0.098 | 0.2751 | 126 |
| **Uganda** | | | | |
| OC43-NP | 4.7 | -0.039 | 0.5699 | 213 |
| X229E- NP | 7.5 | 0.3551 | <0.001 | 213 |
| HKU1- NP | 12.7 | 0.0434 | 0.5285 | 213 |
| NL63 – S1* | 13.2 | 0.0419 | 0.5431 | 213 |
| MERS-NP | 14.6 | -0.0481 | 0.4850 | 213 |

*corr coef is against SARS-CoV-2 RBD domain as the NL63 antigen was the S1 protein

**Suppl. Table 4. A sensitivity analysis to assess if SARS-CoV-2 seroprevalence (RBD/NP IgG/M) differs in samples with evidence of high MFI to other HCoVs[1]**

[1] In the table we defined the HCoV 'seropositive' column as those with IgG MFI units 3 standard deviations above the mean of European negative controls. The data on other HCoVs are based on IgG to OC43 NP, HKU1 NP, 229E NP, NL63 S1 protein

a) DRC

| | | OC43 NP IgG | | | Seroprevalence to OC43 | Chi2 p-value |
|---|---|---|---|---|---|---|
| | | (-) | (+) | Total | | |
| SARS2 RBD/NP IgG / IgM | (-) | 56 | 2 | 58 | 3.4% | |
| | (+) | 135 | 3 | 138 | 2.2% | |
| | Total | 191 | 5 | 196 | 2.6% | 0.606 |
| | Seroprevalence to SARS2 | 71% | 60% | | | |

| | | HKU1 NP IgG | | | Seroprevalence to HKU1 | Chi2 p-value |
|---|---|---|---|---|---|---|
| | | (-) | (+) | Total | | |
| SARS2 RBD/NP IgG / IgM | (-) | 51 | 7 | 58 | 12.1% | |
| | (+) | 123 | 15 | 138 | 10.9% | |
| | Total | 174 | 22 | 196 | 11.2% | |
| | seroprevalence to SARS2 | 71% | 68% | | | 0.808 |

| | | 229E NP IgG | | | Seroprevalence to 229E | Chi2 p-value |
|---|---|---|---|---|---|---|
| | | (-) | (+) | Total | | |
| Luminex RBD/NP IgG / IgM | (-) | 57 | 1 | 58 | 1.7% | |
| | (+) | 134 | 4 | 138 | 2.9% | |
| | Total | 191 | 5 | 196 | 2.6% | |
| | seroprevalence to SARS2 | 70% | 80% | | | 0.634 |

| | | NL63 S1 IgG | | | Seroprevalence to NL63 | Chi2 p-value |
|---|---|---|---|---|---|---|
| | | (-) | (+) | Total | | |
| Luminex RBD/NP IgG / IgM | (-) | 55 | 3 | 58 | 5.2% | |
| | (+) | 132 | 6 | 138 | 4.3% | |
| | Total | 187 | 9 | 196 | 4.6% | |
| | seroprevalence to SARS2 | 71% | 67% | | | 0.801 |

b) Sierra Leone

| | | OC43 NP IgG | | | Seroprevalence to OC43 | Chi2 p-value |
|---|---|---|---|---|---|---|
| | | (-) | (+) | Total | | |
| SARS2 RBD/NP IgG / IgM | (-) | 10 | 4 | 14 | 28.6% | |
| | (+) | 92 | 20 | 112 | 17.9% | |
| | Total | 102 | 24 | 126 | 19.0% | |
| | Seroprevalence to SARS2 | 90% | 83% | | | 0.336 |

| | | HKU1 NP IgG | | | Seroprevalence to HKU1 | Chi2 p-value |
|---|---|---|---|---|---|---|
| | | (-) | (+) | Total | | |
| SARS2 RBD/NP IgG / IgM | (-) | 10 | 4 | 14 | 28.6% | |
| | (+) | 87 | 25 | 112 | 22.3% | |
| | Total | 97 | 29 | 126 | 23.0% | |
| | Seroprevalence to SARS2 | 90% | 86% | | | 0.600 |

| | | 229E NP IgG | | | Seroprevalence to 229E | Chi2 p-value |
|---|---|---|---|---|---|---|
| | | (-) | (+) | Total | | |
| SARS2 RBD/NP IgG / IgM | (-) | 13 | 1 | 14 | 7.1% | |
| | (+) | 109 | 3 | 112 | 2.7% | |
| | Total | 122 | 4 | 126 | 3.2% | |
| | Seroprevalence to SARS2 | 89% | 75% | | | 0.369 |

| | | NL63 S1 IgG | | | Seroprevalence to NL63 | Chi2 p-value |
|---|---|---|---|---|---|---|
| | | (-) | (+) | Total | | |
| SARS2 RBD/NP IgG / IgM | (-) | 14 | 0 | 14 | 0.0% | |
| | (+) | 102 | 10 | 112 | 8.9% | |
| | Total | 116 | 10 | 126 | 7.9% | |
| | Seroprevalence to SARS2 | 88% | 100% | | | 0.244 |

c) Uganda among unvaccinated at baseline only

| | | OC43 NP IgG | | | Seroprevalence to OC43 | Chi2 p-value |
|---|---|---|---|---|---|---|
| | | (-) | (+) | Total | | |
| SARS2 RBD/NP IgG / IgM | (-) | 21 | 2 | 23 | 8.7% | |
| | (+) | 182 | 8 | 190 | 4.2% | |
| | Total | 203 | 10 | 213 | 4.7% | |
| | seroprevalence to SARS2 | 90% | 80% | | | 0.9224 |

| | | HKU1 NP IgG | | | Seroprevalence to HKU1 | Chi2 p-value |
|---|---|---|---|---|---|---|
| | | (-) | (+) | Total | | |
| SARS2 RBD/NP IgG / IgM | (-) | 20 | 3 | 23 | 13.0% | |
| | (+) | 166 | 24 | 190 | 12.6% | |
| | Total | 186 | 27 | 213 | 12.7% | |
| | seroprevalence to SARS2 | 89% | 89% | | | 0.003 |

| | | 229E NP IgG | | | Seroprevalence to HKU1 | Chi2 p-value |
|---|---|---|---|---|---|---|
| | | (-) | (+) | Total | | |
| SARS2 RBD/NP IgG / IgM | (-) | 22 | 1 | 23 | 4.4% | |
| | (+) | 175 | 15 | 190 | 7.9% | |
| | Total | 197 | 16 | 213 | 7.5% | |
| | seroprevalence to SARS2 | 89% | 94% | | | 0.372 |

| | | NL63 S1 IgG | | | Seroprevalence to NL63 | Chi2 p-value |
|---|---|---|---|---|---|---|
| | | (-) | (+) | Total | | |
| SARS2 RBD/NP IgG / IgM | (-) | 20 | 3 | 23 | 13.0% | |
| | (+) | 165 | 25 | 190 | 13.2% | |
| | Total | 185 | 28 | 213 | 13.2% | |
| | seroprevalence to SARS2 | 89% | 89% | | | 0.0002 |

**Suppl. Table 5. Factors associated with remaining persistently negative throughout follow up in the DRC**

| DRC | Control | | Persistently negative | | Total | OR | 95%CI | LRT p-value |
|---|---|---|---|---|---|---|---|---|
| | N | % (col) | N | % (col) | | | | |
| **All** | 163 | 87.6 | 23 | 12.4 | 186 | | | |
| **Age group** | | | | | | | | |
| <30 yrs | 41 | 25.2 | 7 | 30.4 | 48 | 1 | | 0.848 |
| 30-45 yrs | 72 | 44.2 | 9 | 39.1 | 81 | 0.73 | 0.25-2.11 | |
| >45yrs | 50 | 30.7 | 7 | 30.4 | 57 | 0.82 | 0.27-2.53 | |
| **Sex** | | | | | | | | |
| Male | 62 | 38.0 | 18 | 78.3 | 80 | 5.87 | 2.07-16.6 | <0.001 |
| Female | 101 | 62.0 | 5 | 21.7 | 106 | 1 | | |
| **Role [2]** | | | | | | | | |
| Doctor, CO, nurse | 135 | 82.8 | 19 | 82.6 | 154 | 1 | | 0.980 |
| Support staff [4] | 28 | 17.2 | 4 | 17.4 | 32 | 1.01 | 0.57-1.79 | |
| **Highest level of schooling** | | | | | | | | |
| None [7] | | | | | | ~ | | |
| Complete primary | 1 | 0.6 | 0 | 0 | 1 | ~ | | ~ |
| Incomplete secondary | 12 | 7.4 | 2 | 8.7 | 14 | 1.18 | 0.25-5.66 | |
| Complete secondary & above | 149 | 92.0 | 21 | 91.3 | 170 | 1 | | |
| **Smoke once/ week or more** | | | | | | | | |
| No | 162 | 99.4 | 23 | 100 | 185 | ~ | | |
| Yes | 0 | 0 | 0 | 0 | 0 | ~ | | |
| **MUAC** | | | | | | | | |
| Underweight (<24cm) | 13 | 8.0 | 4 | 17.4 | 17 | 2.19 | 0.64-7.55 | 0.287 |
| Normal | 114 | 69.9 | 16 | 69.6 | 130 | 1 | | |
| Obese (>31cm) | 36 | 22.1 | 3 | 13.0 | 39 | 0.59 | 0.16-2.15 | |
| **Known contact with a confirmed COVID-19 case** | | | | | | | | |
| Yes | 42 | 25.8 | 5 | 21.7 | 47 | 1 | | 0.673 |
| No | 121 | 74.2 | 18 | 78.3 | 139 | 1.25 | 0.44-3.57 | |
| **Use of a mask at work** | | | | | | | | |
| None of the time | | | | | | | | |
| Half the time | 27 | 16.6 | 5 | 21.7 | 32 | 1.38 | 0.44-4.33 | 0.834 |
| Most but not all the time | 82 | 50.3 | 11 | 47.8 | 93 | 1 | | |
| All the time | 54 | 33.1 | 7 | 30.4 | 61 | 0.97 | 0.35-2.65 | |
| **Known Pre-existing conditions** | | | | | | | | |
| Yes | 29 | 17.8 | 6 | 26.1 | 35 | 1.63 | 0.59-4.49 | 0.36 |
| No | 134 | 82.2 | 17 | 73.9 | 151 | 1 | | |
| **COVID-19 vaccination** | | | | | | | | |
| Yes | 0 | 0 | 0 | 0 | 0 | | | |
| No | 163 | 100 | 23 | 100 | 186 | | | |

**Suppl. Figure. 4. HCoV IgG MFI units at baseline, comparing those who were seropositive to SARS-CoV-2 at least once during the study ('0' below, n=163) with those remained persistently seronegative to SARS-CoV-2 IgG/M to RBD/ N ('1' below, n=23), in Goma.** P-values are for a Wilcoxon rank-sum test

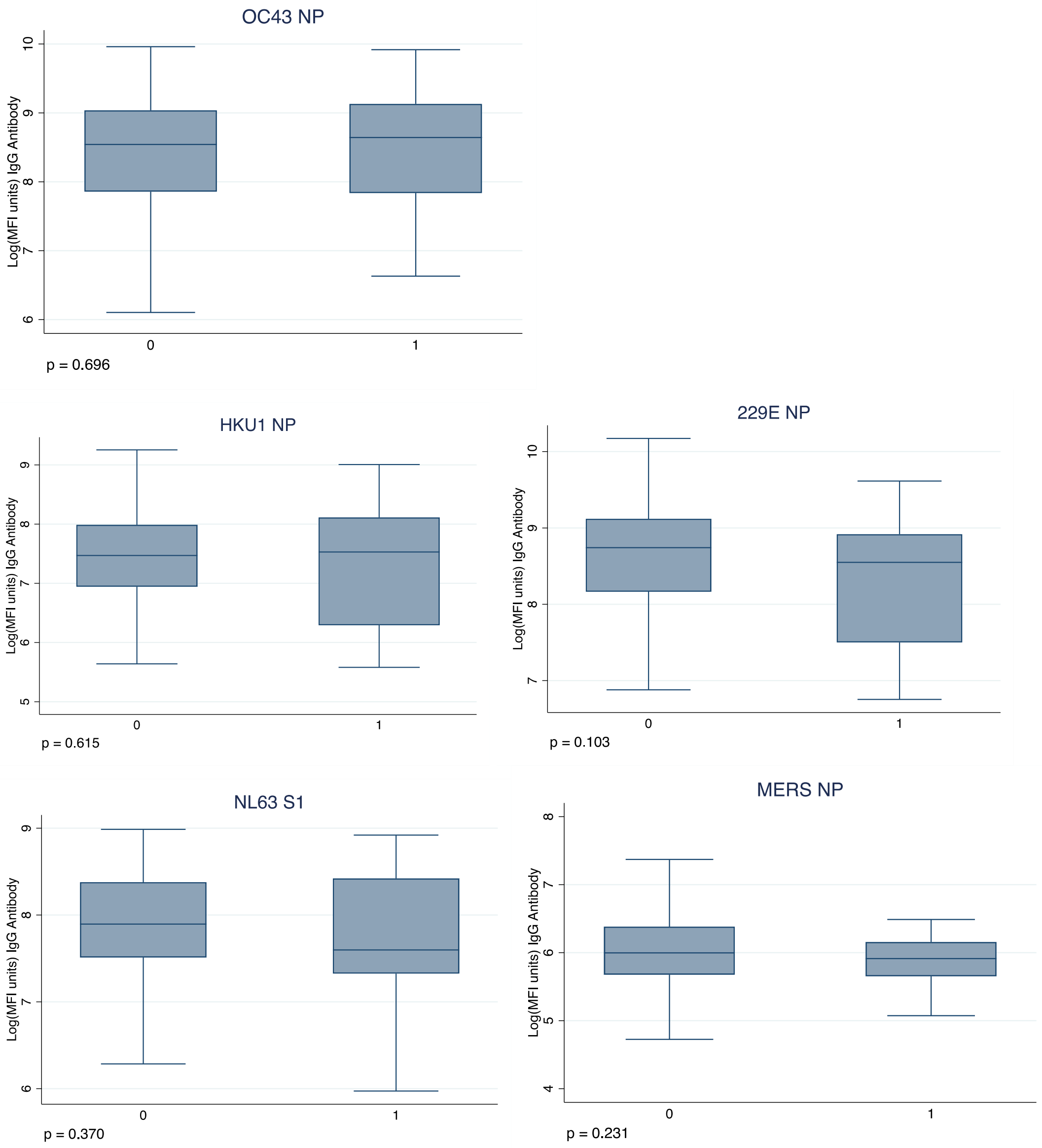

**Suppl. Table 6. Seroprevalence of SARS-CoV-2 in vaccinated**[1]

| Masaka, Uganda | Among those vaccinated before D0 | | | | | | | | |
|---|---|---|---|---|---|---|---|---|---|
| | D0 | | | M2 | | | M4 | | |
| | N (Total) | n (pos) | % | N (Total) | n (pos) | % | N (Total) | n (pos) | % |
| SARS2 RBD IgG | 35 | 30 | 85.7 | 35 | 32 | 91.4 | 34 | 32 | 94.1 |
| SARS2 NP IgG | 35 | 23 | 65.7 | 35 | 22 | 62.9 | 34 | 23 | 67.6 |
| SARS2 RBD/NP IgG | 35 | 31 | 88.6 | 35 | 33 | 94.3 | 34 | 32 | 94.1 |
| SARS2 RBD/NP IgG/IgM | 35 | 33 | 94.3 | 35 | 33 | 94.3 | 34 | 34 | 100.0 |

| Masaka, Uganda | Among those vaccinated between M0 and M2 | | | | | | Among those vaccinated between M2 and M4 | | |
|---|---|---|---|---|---|---|---|---|---|
| | M2 | | | M4 | | | M4 | | |
| | N (Total) | n (pos) | % | N (Total) | n (pos) | % | N (Total) | n (pos) | % |
| SARS2 RBD IgG | 54 | 47 | 87.0 | 51 | 46 | 90.2 | 41 | 33 | 80.5 |
| SARS2 NP IgG | 54 | 36 | 66.7 | 51 | 33 | 64.7 | 41 | 28 | 68.3 |
| SARS2 RBD/NP IgG | 54 | 49 | 90.7 | 51 | 48 | 94.1 | 41 | 33 | 80.5 |
| SARS2 RBD/NP IgG/IgM | 54 | 49 | 90.7 | 51 | 50 | 98.0 | 41 | 36 | 87.8 |

| Kambia, Sierra Leone | Among those vaccinated between M0 and M2 | | | | | | Among those vaccinated between M2 and M4 | | |
|---|---|---|---|---|---|---|---|---|---|
| | M2 | | | M4 | | | M4 | | |
| | N (Total) | n (pos) | % | N (Total) | n (pos) | % | N (Total) | n (pos) | % |
| SARS2 RBD IgG | 40 | 34 | 85 | 40 | 24 | 60 | 12 | 7 | 58.3 |
| SARS2NP IgG | 40 | 27 | 67.5 | 40 | 29 | 72.5 | 12 | 5 | 41.7 |
| SARS2 RBD/NP IgG | 40 | 35 | 87.5 | 40 | 32 | 80 | 12 | 8 | 66.7 |
| SARS2 RBD/NP IgG/IgM | 40 | 37 | 92.5 | 40 | 33 | 82.5 | 12 | 8 | 66.7 |

[1]In DRC: all participants remained unvaccinated for the duration of the study. The tables are based on timing of first dose, some of these participants may have been given a second dose between month 2 and month 4

**Suppl. Figure 5. Trends in IgG MFI to SARS-COV-2 RBD by time since vaccination in Sierra Leone and Uganda.** Curves are estimated using LOWESS, locally weighted smoothing of scatterplot data. Dashed line is the cut off for seropositivity in each setting.

a) Sierra Leone[1]

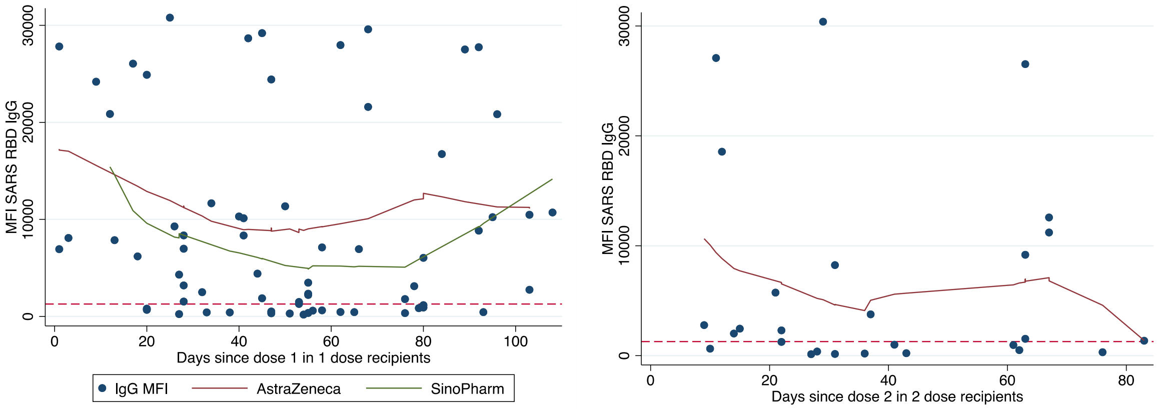

b) Uganda[2]

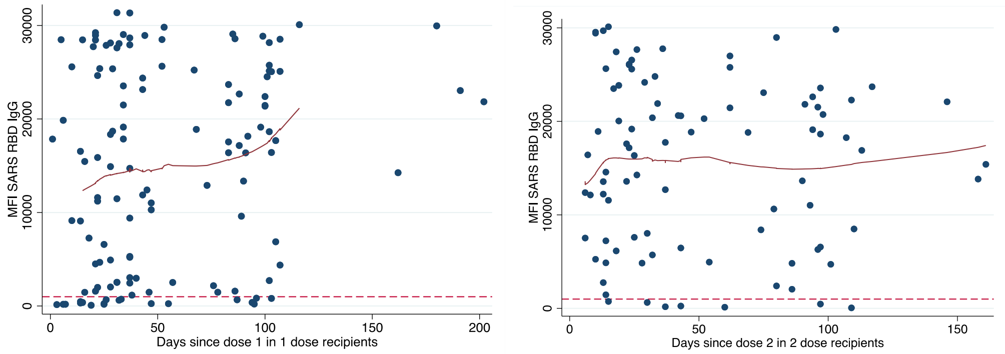

[1] In Sierra Leone, all participants had written records of vaccination and product: AstraZeneca dose 1 recipients n=45; Sinopharm dose 1 recipients n=33. Due to small numbers AstraZeneca and Sinopharm recipients were combined for the dose 2 analysis n=33.

[2] In Uganda 128 participants submitted data on the date of dose 1. Only 14 participants had a written record with product recorded in Uganda, and some of these dates could be mis-remembered as based on recall. A total of 91 participants submitted data on the date of dose 2. Four dates were excluded as outliers when we estimated the trend using LOWESS.

**Suppl. Table 7: Analyses and sample sizes**

| | Research questions | Sample size | | | Refer to table/ figure |
|---|---|---|---|---|---|
| | | DRC | SL | UG | |
| 1 | Among unvaccinated: Prevalence of IgG/IgM to SARS2 as indicator of natural exposure (extent of transmission) | V1=196<br>V2=189<br>V2=189 | V1=126<br>V2=75<br>V3=62 | V1=213<br>V2=141<br>V3=98 | Table 2 |
| 2 | Among unvaccinated: Rate of waning of IgG after natural infection? Restricted to those persistently positive over time to atttempt to restrict to a group with similar time of infection | n=119 | n=41 | n=83 | Table 3, Fig 2 |
| 3 | Among vaccinated: seroprevalence over time among those vaccinated before baseline (group 1), vaccinated between V1-V2 (group 2) and vaccinated between V2 and V3 (group 3) | 0 | Group1=35<br>group2=54<br>group3=41 | Group 1=0<br>Group 2=40<br>Group 3= 12 | Table 4 |
| 4 | Among vaccinated: MFI units by time since vaccination (waning?) and prevalence of hybrid immunity | 0 | 78 | 128 | Supplementary information |
| 5 | Among vaccinated: does evidence of prior natural exposure affect vaccine responses? | 0 | seronegative at baseline=4 vs. seropositive at baseline=44 | seronegative at baseline=12 vs seropositive at baseline=96 | Figure 3 |
| 6 | Among unvaccinated: Do responses to HCoVs correlate with SARS2 and do they interfere with seroprevalence estimates? | V1=196<br>V2=189<br>V2=189 | V1=126<br>V2=75<br>V3=62 | V1=213<br>V2=141<br>V3=98 | Supplementary information |
| 7 | Among unvaccinated: Does IgG to HCoVs influence subsequent risk of acquisition (among those with no evidence of prior infection ) | Remained uninfected=23 vs infected during follow up=28 | 0 | 0 | Figure 1 |
